# Toward trustworthy clinical AI for obsessive-compulsive disorder: reliability, generalizability, and interpretability of a transformer model across the ENIGMA-OCD consortium

**DOI:** 10.64898/2026.04.24.26351711

**Authors:** Maria Pak, Youngchan Ryu, Sangyoon Bae, Alan Anticevic, Ana Daniela Costa, Anders L. Thorsen, Anouk L. van der Straten, Beatriz Couto, Benedetta Vai, Bjarne Hansen, Carles Soriano-Mas, Chiang-shan R. Li, Chris Vriend, Christine Lochner, Christopher Pittenger, Clara A. Moreau, Daniela Rodriguez-Manrique, Daniela Vecchio, Eiji Shimizu, Emily R. Stern, Emma Muñoz-Moreno, Erika L. Nurmi, Fabrizio Piras, Federica Colombo, Federica Piras, Fern Jaspers-Fayer, Francesco Benedetti, Ganesan Venkatasubramanian, Goi Khia Eng, H. Blair Simpson, Hanyang Ruan, Hao Hu, Hein J.F. van Marle, Hirofumi Tomiyama, Ignacio Martínez-Zalacaín, Jamie Feusner, Janardhanan C. Narayanaswamy, Je-Yeon Yun, Joao R. Sato, Jonathan Ipser, Jose C. Pariente, Jose M. Menchón, Joseph O’Neill, Jun Soo Kwon, Kathrin Koch, Kristen Hagen, Lea Backhausen, Lea Waller, Luisa Lazaro, Marcelo C. Batistuzzo, Marcelo Q. Hoexter, Maria Picó-Pérez, Minah Kim, Nadza Dzinalija, Nicole Beyer, Nora C. Vetter, Patricia Gruner, Pedro Morgado, Philip R. Szeszko, Pino Alonso, Qing Zhao, Rachel Marsh, S. Evelyn Stewart, Sara Bertolín, Silvia Brem, Sophia I. Thomopoulos, Srinivas Balachander, Susanne Walitza, Tokiko Yoshida, Tomohiro Nakao, Venkataram Shivakumar, Wieke van Leeuwen, Y.C. Janardhan Reddy, Yoshinari Abe, Yoshiyuki Hirano, Youngsun Cho, Ysbrand D. van der Werf, Yuki Ikemizu, Yuki Sakai, Zhen Wang, ENIGMA-OCD Working Group, Paul M. Thompson, Willem Bruin, Guido van Wingen, Dan J. Stein, Odile A. van den Heuvel, Jiook Cha, Ana Vígil-Perez, Anri Watanabe, Beatrice Bravi, Blanca García-Delgar, Cinto Segalàs, Dick J. Veltman, Eugenie Choe, Eva Real, Freda Scheffler, Gerd Kvale, Hitomi Kitagawa, Jin Narumoto, Jinsong Tang, John C. Piacentini, Joyce Guo, Kayleigh Beukes, Kei Yamada, Koji Matsumoto, Laurens van de Mortel, Leila Darwich, Mafalda M. Sousa, Marta Subirà, Minji Ha, Nuno Sousa, Olga Therese Ousdal, Pedro S. Moreira, Rachael Grazioplene, Renata Melo, Roseli G. Shavitt, Sara Dallaspezia, Takashi Nakamae, Toshikazu Ikuta, Veit Roessner, Yuko Isobe

## Abstract

**Background:** Studies applying machine learning to obsessive-compulsive disorder (OCD) typically report accuracy in homogeneous samples but rarely assess model reliability, generalizability, and interpretability needed for clinical use.

**Methods:** We applied a transformer-based deep learning model, the Multi-Band Brain Net, to the ENIGMA-OCD cohort – the largest available resting-state functional magnetic resonance imaging (rs-fMRI) dataset in OCD with 1,706 participants (869 cases with OCD, 837 controls) across 23 sites worldwide. We evaluated model reliability by calculating calibration – the model’s ability to “know what it doesn’t know”. We assessed generalizability using leave-one-site-out validation to test performance on unseen sites with different scanners, acquisition protocols, and patient populations. Finally, we examined interpretability by analyzing model attention weights to identify the neural connectivity patterns that influence model predictions.

**Results:** The model achieved modest but competitive classification performance (AUROC = .653 ± .039). Crucially, while large-scale pretraining on the UK Biobank (N = 40,783) did not boost accuracy, it significantly enhanced model calibration by reducing overconfident predictions. Leave-one-site-out validation showed a generalization gap across sites (AUROC = .427-.819). Pretraining did not close this gap but removed scanner manufacturer bias. Finally, attention-based mapping identified biologically plausible patterns of widespread hypoconnectivity in OCD relative to healthy controls, particularly in low-frequency bands involving the default mode, salience, and somatomotor networks. These findings aligned with known OCD neurobiology.

**Conclusions:** This study provides a framework for developing more reliable and trustworthy clinical artificial intelligence for OCD.

## Introduction

Obsessive-compulsive disorder (OCD) is a chronic and debilitating illness affecting 2-3% of the global population^1^. The disorder is associated with substantial functional impairment, reduced quality of life, and a considerable societal and economic burden^1^. In clinical practice, diagnosis relies primarily on symptom-based criteria assessed through structured interviews and clinician judgment^2,3^. While these approaches are the gold diagnostic standard, the heterogeneity of the disorder and limited inter-rater reliability create a need for objective tools to complement clinical assessment^4,5^. Converging neurobiological models of OCD implicate alterations in cortico-striato-thalamo-cortical circuits, extending to spatial-attentional and executive networks^6,7^, suggesting that the disorder is associated with measurable changes in brain structure and function^1^. These advances imply that neuroimaging may contribute objective biological markers that inform clinical evaluation. Rather than replacing clinical judgment, such tools could translate neural data into stable and reproducible markers that provide individualized biological insights for clinical decision support^8,9^.

Recent neuroimaging studies suggest that OCD involves broad disruptions across large-scale brain networks^10^. Large collaborative efforts such as the ENIGMA-OCD consortium have reported alterations in functional connectivity involving the default mode, salience, and sensorimotor networks^11,12^. These networks support processes central to OCD symptomatology, including internally directed thought, salience detection, cognitive control, and the integration of sensory and motor signals that guide habitual and repetitive behaviors^13^. Yet most neuroimaging findings are derived from group-level analyses, and high heterogeneity of brain patterns at the individual level limits their clinical utility. Machine learning approaches have therefore been increasingly applied to translate these complex neuroimaging signals into individual-level classification models^9^.

Previous studies applied machine learning methods to classify patients with OCD from healthy controls using neuroimaging data. A recent meta-analysis reported classification accuracies ranging from 66% to 100% across studies using structural and functional magnetic resonance imaging (MRI)^4^. Most studies rely on conventional machine learning models such as support vector machines^4^. Some studies have also explored features derived from specific frequency bands of the resting-state functional MRI (rs-fMRI) signal. These studies show that different frequency components of the blood-oxygen-level-dependent (BOLD) signal may contain distinct information about brain activity, helping models identify disease-related patterns more effectively^14,15^. However, most of these studies are based on relatively small and homogeneous datasets, typically consisting of fewer than 100 participants from a single research site^4^. Models trained in such settings may capture patterns specific to a particular scanner, acquisition protocol, or study population, rather than generalizable disease-related signals.

As a result, models optimized within a single dataset may fail when applied to independent cohorts – a challenge that is particularly pronounced in large multi-site consortia such as ENIGMA-OCD, where differences in scanners, acquisition protocols, and patient populations create systematic domain shifts^16,17^. Indeed, ENIGMA-OCD analyses have reported only moderate classification performance^11,18,19^ in sharp contrast to the high accuracies seen in single-site studies, suggesting that those results may largely reflect overfitting to site-specific signals. This phenomenon – commonly referred to as the generalization problem^20^ – underscores the difficulty of translating machine learning findings from homogeneous research settings to real-world clinical populations.

However, poor generalization is only one obstacle to clinical deployment. In high-stakes applications such as clinical decision-making in psychiatry, models must also be reliable and interpretable, in addition to maintaining performance across independent sites, scanners, and patient populations. Reliability refers to the ability of a model to quantify prediction uncertainty for individual patients – analogous to a confidence interval around each prediction – rather than only reporting overall accuracy across the dataset^21–23^. Without such estimates, artificial intelligence (AI) systems produce a single prediction score with no indication of its stability for a given individual^22,24^. Interpretability is also essential, ensuring that predictions rely on biologically plausible neural signals rather than site-specific artifacts or noise^4^. Despite their importance for clinical deployment, few neuroimaging studies evaluate reliability, generalizability, and interpretability simultaneously.

Recent advances in deep learning suggest several methodological strategies that may help address these challenges. One promising approach involves the transformer architecture^25^ – a type of neural network originally developed for language processing that has recently been applied to brain imaging data^9^. A key feature of transformers is an attention mechanism that identifies the most informative parts of the brain signal^25^. This mechanism may improve interpretability by highlighting the functional connectivity patterns that contribute most strongly to the model’s predictions^26^. Models based on the transformer architecture also support methods for estimating the reliability of individual predictions, allowing the model to indicate when a prediction for a particular patient may be uncertain^22^. Another important capability is pretraining on large neuroimaging datasets before application to smaller clinical samples, which may help models better handle the variability typically seen in clinical datasets^27,28^. However, it remains unclear whether the transformer architecture and large-scale pretraining truly improve model classification performance, reliability, generalizability, and interpretability when applied to heterogeneous psychiatric datasets such as ENIGMA-OCD.

To address these gaps, we apply a transformer architecture – specifically, the Multi-Band Brain Net (MBBN)^29^ – to the largest available multi-site cohort of 869 cases with OCD and 837 healthy controls. The MBBN decomposes rs-fMRI time series into multiple frequency bands, capturing complementary neural dynamics across frequency scales^14,15^. The model learns patterns of temporal dynamics and functional connectivity that distinguish individuals with OCD from healthy controls. We compared its performance with several conventional machine learning and deep learning approaches, including support vector machines, gradient-boosted trees, and alternative transformer architectures. We further examine whether large-scale self-supervised pretraining on 40,783 individuals from the UK Biobank improves model performance and robustness to scanner-related variability. Beyond classification accuracy, we assess the model’s prediction uncertainty, evaluate its ability to generalize across data sites, and identify the functional connectivity patterns contributing to model decisions. Together, this framework aims to move psychiatric AI beyond raw classification toward models that are reliable, generalizable, and interpretable – key requirements for future clinical translation.

## Results

### 1. Study participants and clinical characteristics

To address the limitations of small-scale neuroimaging studies, we analyzed a global dataset from the ENIGMA-OCD Working Group comprising 1,706 participants (869 OCD patients and 837 healthy controls, HCs) across 23 international sites worldwide (Supplementary Table S1, Supplementary Figure S1). This cohort represents the largest resting-state functional magnetic resonance imaging (rs-fMRI) OCD study to date, providing the statistical power necessary for robust deep learning applications. To ensure compatibility between the ENIGMA-OCD dataset and the UK Biobank dataset used for model pretraining, the analysis was restricted to cortical regions from the Schaefer atlas and standardized time-series lengths across datasets (see Methods).

The groups were well-balanced for age (OCD: 29.63 ± 10.98 years; HC: 28.55 ± 9.70 years; *p* = .092; Supplementary Figure S2). There was a significant difference in sex distribution (OCD: 45.2% male; HC: 50.9%; *p* = .019; Table 1), but the corresponding effect size was small (*phi* = 0.057). Those with OCD exhibited moderate-to-severe symptom severity (mean Y-BOCS score: 24.34 ± 6.68; Table 1), with high prevalences of aggressive or checking (56.5%) and contamination or cleaning (49.5%) symptoms (Supplementary Table S2). Together, the large sample size and multi-site design capture substantial clinical and imaging heterogeneity, providing a realistic setting for evaluating machine learning models for OCD.

**Table 1.** Demographics and clinical characteristics. Continuous variables are presented as mean ± standard deviation. Statistical tests: Mann-Whitney U tests for continuous variables (age, education); chi-square test for categorical variables (sex). Effect sizes: rank-biserial correlation (*r*) for Mann-Whitney U tests; phi coefficient (*φ*) for chi-square test. **Abbreviations:** OCD: obsessive-compulsive disorder, HC: healthy controls, Y-BOCS: Yale-Brown obsessive-compulsive scale.

| Characteristic | OCD | HC | Missing OCD | Missing HC | *p* | Effect size |
| --- | --- | --- | --- | --- | --- | --- |
| **N** | 869 | 837 | - | - | - | - |
| **Age, years** | 29.63 ± 10.98 | 28.55 ± 9.70 | 0 (0.0%) | 0 (0.0%) | .092 | 0.047 |
| **Sex, n (%)**<br>[Male / Female] | 393 (45.2)<br>/ 476 (54.8) | 426 (50.9) /<br>411 (49.1) | 0 (0.0%) | 0 (0.0%) | .019 | 0.057 |
| **Education, years** | 13.18 ± 3.72 | 15.04 ± 3.57 | 30 (3.5%) | 121 (14.5%) | < .001 | 0.304 |
| **Y-BOCS total score** | 24.34 ± 6.68 | - | 13 (1.5%) | - | - | - |
| **Age of onset, n (%)**<br>[Child- / Adult-onset] | 424 (48.8)<br>/ 329 (37.9) | - | 116 (13.3%) | - | - | - |
| **Medication use at time of scan, n (%)**<br>[Yes / No] | 447 (51.4)<br>/ 415 (47.8) | - | 7 (0.8%) | - | - | - |

### 2. Multi-band model achieves competitive classification performance

We evaluated the performance of the Multi-Band Brain Net (MBBN) model (Figure 1) against several established machine learning and deep learning approaches commonly used in neuroimaging-based classification, including XGBoost, support vector machines, the brain network transformer (a previously proposed transformer architecture designed for brain connectivity data), and the foundation model BrainLM (a large pretrained neural network trained on large-scale neuroimaging data).

**Figure 1.**
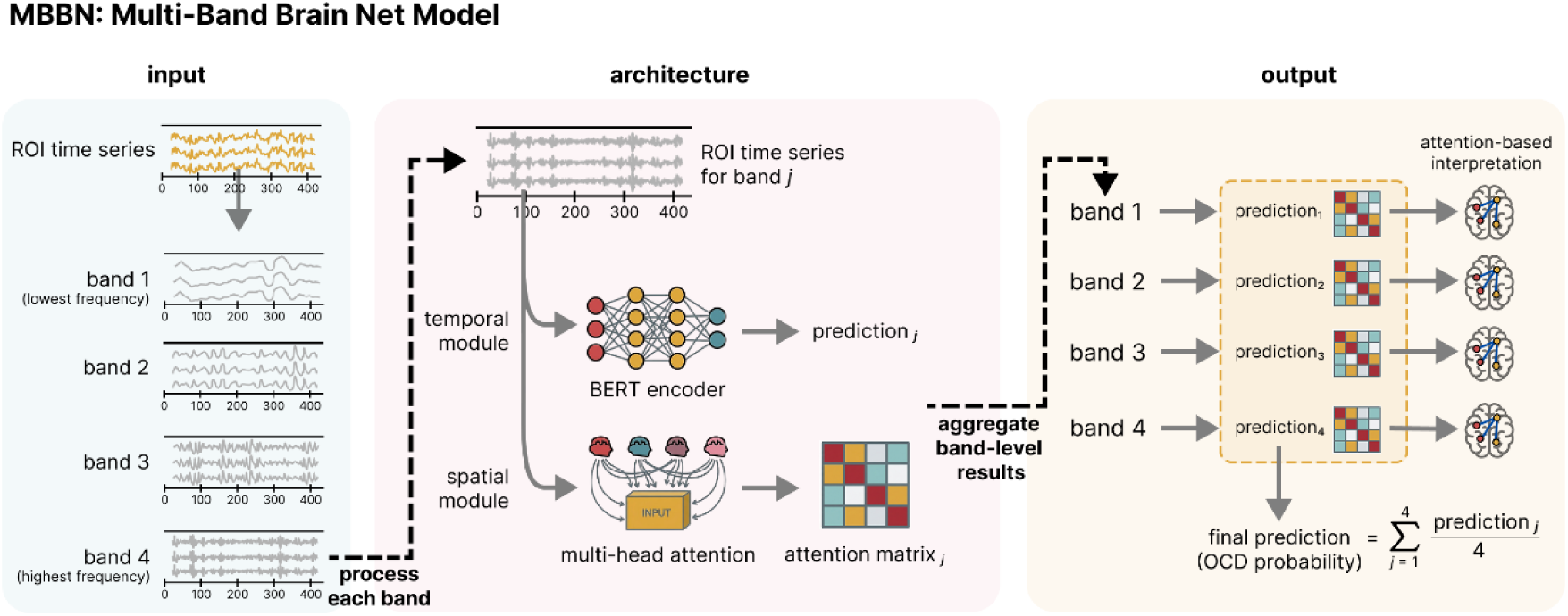
MBBN model architecture for OCD classification. The pipeline processes resting-state fMRI data from the ENIGMA-OCD dataset. **Input:** time series from 304 ROIs are decomposed into four frequency bands via variational mode decomposition (VMD). **Architecture:** each band passes through two parallel modules. The temporal module uses a BERT encoder with shared weights across frequency bands, producing a prediction score for each band. The spatial module uses multi-head attention with frequency-specific weights to learn functional connectivity patterns, producing an attention matrix for each band. **Output:** the model averages band-specific logits for final prediction and extracts attention matrices for frequency-specific interpretability. **Abbreviations:** OCD: obsessive-compulsive disorder, ROI: region of interest, BERT: bidirectional encoder representations from transformers.

The MBBN model trained from scratch achieved the highest mean area under the receiver operating characteristic curve (AUROC = .653 ± .039, standard deviation; 95% corrected CI [.601, .704]) in a large multi-site setting across 10 independent train-validation-test data splits (see Methods), significantly exceeding chance-level classification (*p* < .001) (Figure 2A). Statistical comparisons showed that the MBBN significantly outperformed BrainLM (AUROC = .568 ± .038; *p*_FDR_ = .035), while the difference with XGBoost approached but did not survive multiple comparison correction (AUROC = .593 ± .031; *p*_FDR_ = .065). Performance was comparable to the support vector machine (AUROC = .642 ± .021; *p*_FDR_ = .708) and brain network transformer (BNT; AUROC = .648 ± .041; *p*_FDR_ = .911). Detailed model performance is reported in Supplementary Table S4.

**Figure 2.**
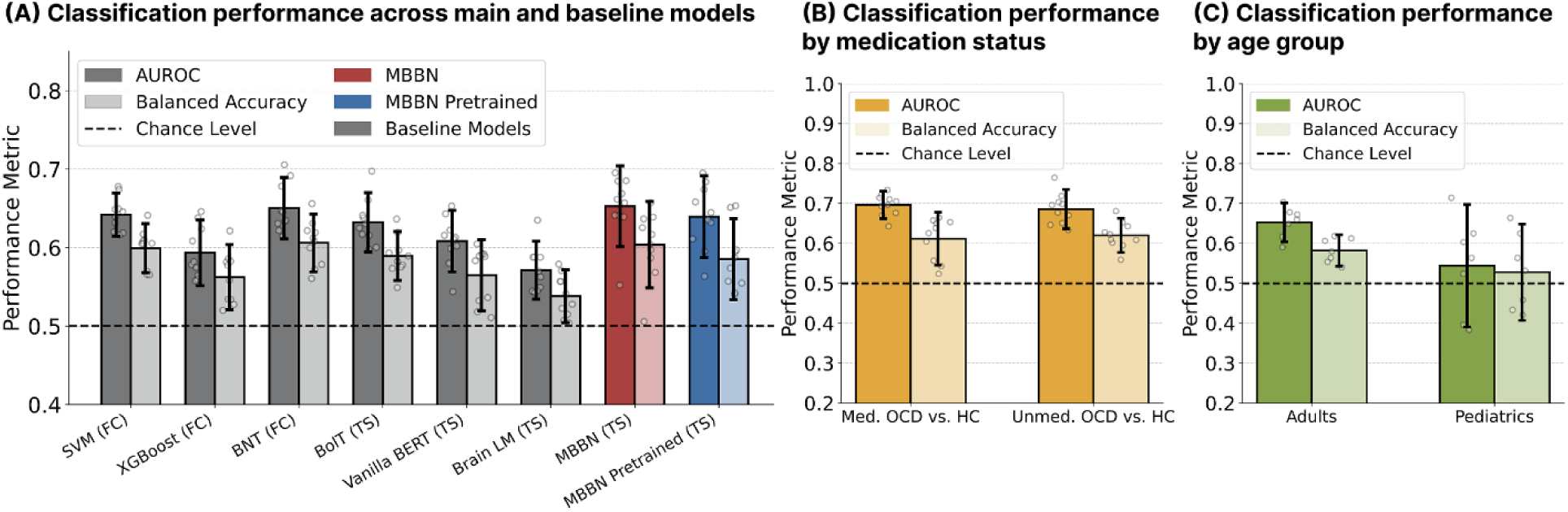
Model performance comparison across baseline models and clinical subgroups. Error bars represent 95% confidence intervals across 10 random splits, dashed lines represent the chance level. **(A)** The from-scratch MBBN model (red) achieved the highest AUROC (.653), significantly outperforming Brain LM and showing a trend toward outperforming XGBoost that did not survive FDR correction. **(B)** From-scratch MBBN performance remained stable across medicated (AUROC = .697) and unmedicated (.686) subgroups. **(C)** Performance was stable in adults (.653) but lower in pediatric populations (.544), likely due to smaller sample sizes. **Abbreviations:** AUROC: area under the receiver operating characteristic curve, FC: functional connectivity, TS: time series, SVM: support vector machine, XGBoost: extreme gradient boosting, BNT: brain network transformer, BERT: bidirectional encoder representations from transformers, BolT: blood-oxygen-level-dependent transformer.

Our model demonstrated consistent classification performance across clinical subgroups, including medicated patients (AUROC = .697 ± .026; 447 OCD vs 837 HC), unmedicated patients (AUROC = .686 ± .037; 415 OCD vs 837 HC), and the total adult sample (AUROC = .653 ± .036; 760 OCD vs 754 HC) (Figure 2B-C, Supplementary Table S11). Performance was substantially lower and showed greater variability across splits in the pediatric subgroup (AUROC = .544 ± .112; 109 OCD vs 83 HC).

We further evaluated whether large-scale self-supervised pretraining on the UK Biobank (N = 40,783) improved classification performance. The pretrained MBBN achieved slightly lower AUROC (.639 ± .039) than the model trained from scratch (.653 ± .039), but the difference was not statistically significant (*p*_FDR_ = .663). Lastly, our control analysis demonstrated that four frequency bands yielded the best classification results compared to configurations using one to five bands (Supplementary Table S6), aligning with the previous results on rs-fMRI signal frequency decomposition^30^.

### 3. Pretraining enhances model calibration and reliability

The model initialized with large-scale pretraining on the UK Biobank showed significantly lower expected calibration error than the same architecture trained from scratch (*p* < .001), with a non-significant trend toward lower maximum calibration error (*p* = .067) (Figure 3A). This resulted in confidence scores that more accurately reflected the probability of predicting a correct diagnosis. Specifically, the pretrained model produced fewer overconfident predictions in the 90-100% confidence range compared to the from-scratch model (Figure 3B).

**Figure 3.**
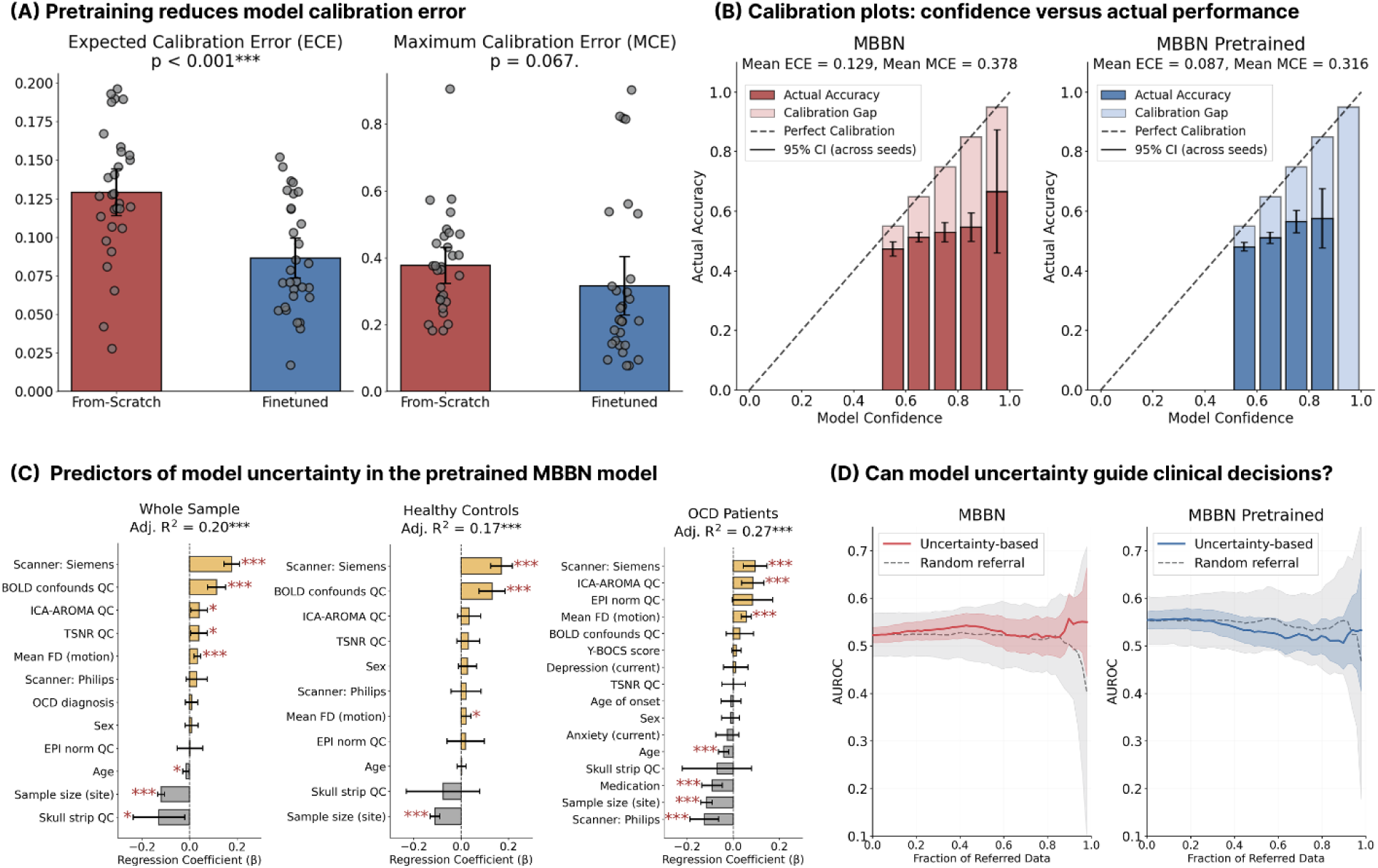
Impact of pretraining on model calibration and clinical utility of uncertainty. **(A–B)** The pretrained MBBN model shows significantly lower expected calibration error (ECE; *p* < .001) and a non-significant trend toward lower maximum calibration error (MCE; *p* = .067), with fewer overconfident predictions than the from-scratch model. Error bars represent 95% confidence intervals across 30 random splits. **(C)** Multiple linear regression reveals uncertainty predictors (e.g., data quality, scanner manufacturer, head motion) across whole-sample and subgroup models. Positive coefficients indicate factors associated with higher uncertainty. Quality control (QC) variables are binary-coded quality ratings (0 = good, 1 = uncertain). Significance levels: *p* < .05*, *p* < .01**, *p* < .001***. **(D)** Uncertainty-based referral (red/blue) did not improve AUROC over random referral (gray), suggesting uncertainty does not reliably predict misclassification. **Abbreviations:** MBBN: Multi-Band Brain Net, OCD: obsessive-compulsive disorder, AUROC: area under the receiver operating characteristic curve, BOLD: blood-oxygen-level-dependent, ICA-AROMA: independent component analysis-based automatic removal of motion artifacts, TSNR: temporal signal-to-noise ratio, FD: framewise displacement, EPI: echo planar imaging, Y-BOCS: Yale-Brown obsessive-compulsive scale.

To understand what affects the model’s confidence, we performed multiple linear regression to test whether technical and clinical variables predicted higher or lower uncertainty in the model’s predictions. For the pretrained model, higher uncertainty was systematically associated with Siemens scanners (*β* = 0.177, *p* < .001, reference level: GE), higher head motion (*β* = 0.033, *p* < .001), and poor data quality (e.g., uncertain BOLD confound ratings: *β* = 0.113, *p* < .001). Conversely, lower uncertainty was consistently associated with larger site sample sizes (*β* = - 0.120, *p* < .001), suggesting that the model produces more reliable predictions for larger sites (site sample sizes ranged from 29 to 358 participants) (Figure 3C).

Within the OCD group, medication use was associated with significantly lower uncertainty (*β* = - 0.092, *p* < .001) (Figure 3C, Supplementary Table S13), whereas higher uncertainty was observed in patients with the symmetry or ordering symptom dimension (*p*_FDR_ = .010) and those with comorbid anxiety compared to comorbid depression (*p*_FDR_ = .039) (Supplementary Figure S4B-C). Uncertainty scores did not differ between the OCD and healthy control groups (Supplementary Figure S4A). Despite improved calibration, a decision referral analysis – simulation of the uncertainty-based referral strategy whereby high-uncertainty cases are referred to human experts – did not significantly improve overall classification accuracy (Figure 3D).

### 4. Site-level technical heterogeneity limits model generalization

To evaluate the real-world robustness of our approach, we performed leave-one-site-out cross-validation (LOSO-CV). This analysis revealed a significant “generalization gap”, with from-scratch classification performance varying from AUROC of .427 to .819 across the 23 international sites (Figure 4A). The LOSO performance of the pretrained model was not significantly higher than that of the from-scratch model (*p* = .136) (Figure 4B).

**Figure 4.**
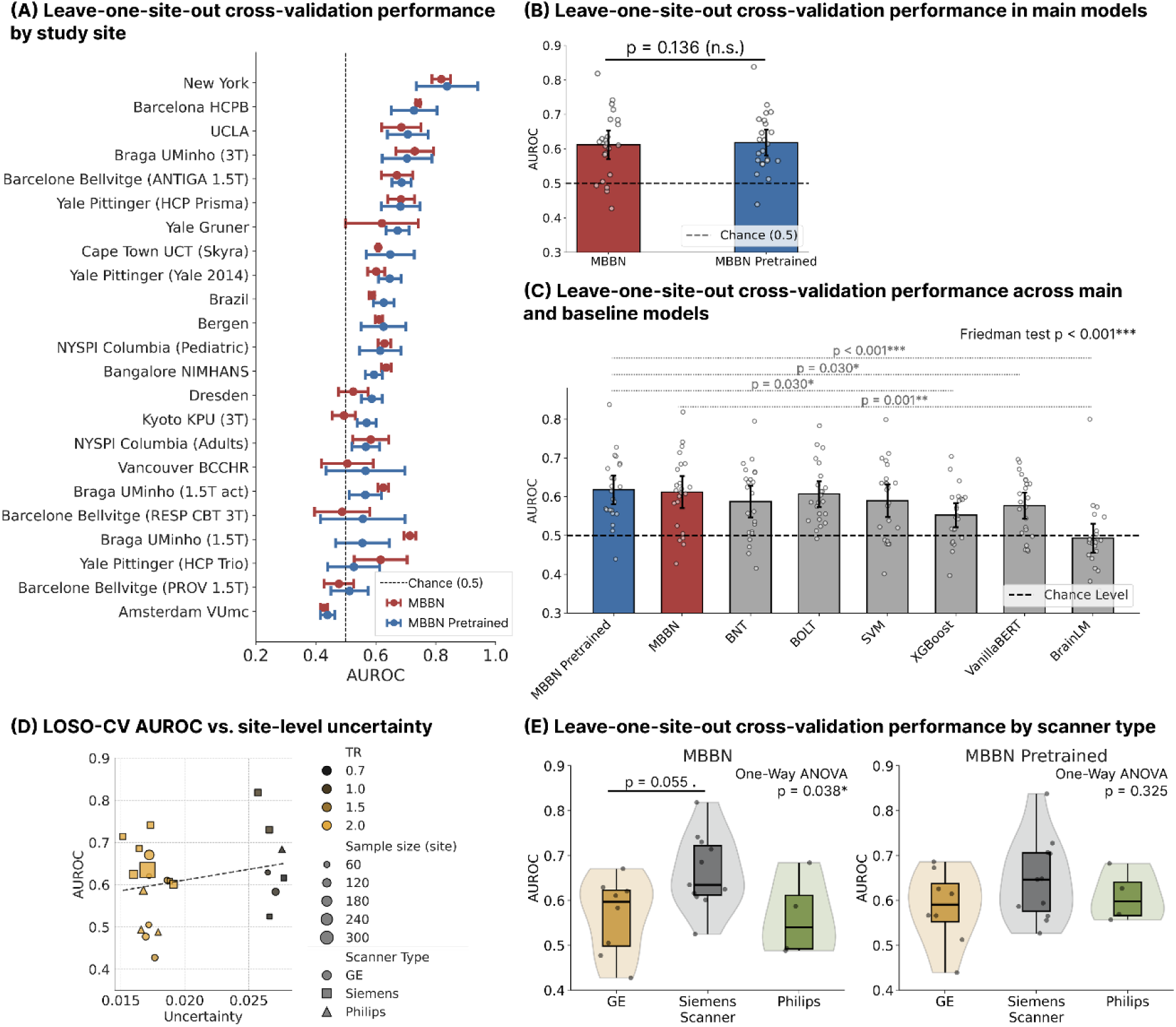
Model generalizability across sites and scanners. Results from leave-one-site-out cross-validation (LOSO-CV) on 23 sites. For each site, AUROC scores were averaged across three independent data splits. **(A)** Performance varies by site; error bars represent standard deviations across three random data splits for each test site. **(B)** Pretraining did not improve model generalizability compared to the from-scratch model (*p* = .136). **(C)** Pretrained MBBN significantly outperformed XGBoost (*p =* .030), vanilla BERT (*p =* .030), and Brain LM (*p <* .001), and matched other baselines. **(D)** Sites cluster by repetition time (TR); higher TR correlates with lower uncertainty. **(E)** Pretraining eliminated scanner-dependent performance biases (*p* = .038 in from-scratch vs. .325 in pretrained), enhancing hardware invariance. **Abbreviations:** MBBN: Multi-Band Brain Net, AUROC: area under the receiver operating characteristic curve, SVM: support vector machine, XGBoost: extreme gradient boosting, BNT: brain network transformer, BERT: bidirectional encoder representations from transformers, BolT: blood-oxygen-level-dependent transformer.

In benchmarking against established models, a Friedman test confirmed significant differences in generalizability across architectures (*χ*^2^ = 34.48, *p* < .001). Post-hoc pairwise comparisons revealed that while both MBBN versions significantly outperformed the Brain LM foundation model (*p* < .001), they achieved performance comparable to traditional benchmarks such as the support vector machine (pretrained MBBN vs. SVM: *p*_FDR_ = .317) and a graph-based brain network transformer (pretrained MBBN vs. BNT: *p*_FDR_ = .317) (Figure 4C).

To quantify the potential influence of site-level variability, we performed the intra-class correlation (ICC) analysis, which revealed that approximately 68% of the variance in functional connectivity data was attributable to site-level factors rather than individual-level differences (Supplementary Figure S5). Qualitative analysis showed two groups of sites with different uncertainty levels associated with repetition time (TR): sites with lower TRs exhibited higher site-level uncertainty scores (Figure 4D). Further investigation into site-level technical factors showed that the from-scratch model’s LOSO performance was significantly influenced by scanner manufacturer (*p* = .038), whereas this effect was not significant in the pretrained model (*p* = .325) (Figure 4E).

### 5. Frequency-specific connectivity patterns learned by the model

To examine which neuroimaging features contributed most strongly to classification, we analyzed attention weights produced by the spatial module of the model. These weights indicate which functional connectivity edges most influenced the model’s predictions at different frequency bands. The data-driven frequency band boundaries were consistent with previous literature^30^: Band 1 (0.004-0.04 Hz), Band 2 (0.04-0.10 Hz), Band 3 (0.10-0.17 Hz), and Band 4 (0.17-0.25 Hz) (Supplementary Table S8).

We found that connectivity patterns derived from the model’s attention weights differed between OCD patients and healthy controls, particularly in the lower frequency bands. Band 1 exhibited the highest density of discriminatory connections (∼18,000), followed by Band 2 (∼6,000), while higher frequency bands contained very few significant connections among brain regions (Figure 5A). Across these bands, the dominant pattern was widespread hypoconnectivity in OCD patients compared with healthy controls. These differences involved connections within and between the default mode, salience, and somatomotor networks (Figure 5).

**Figure 5.**
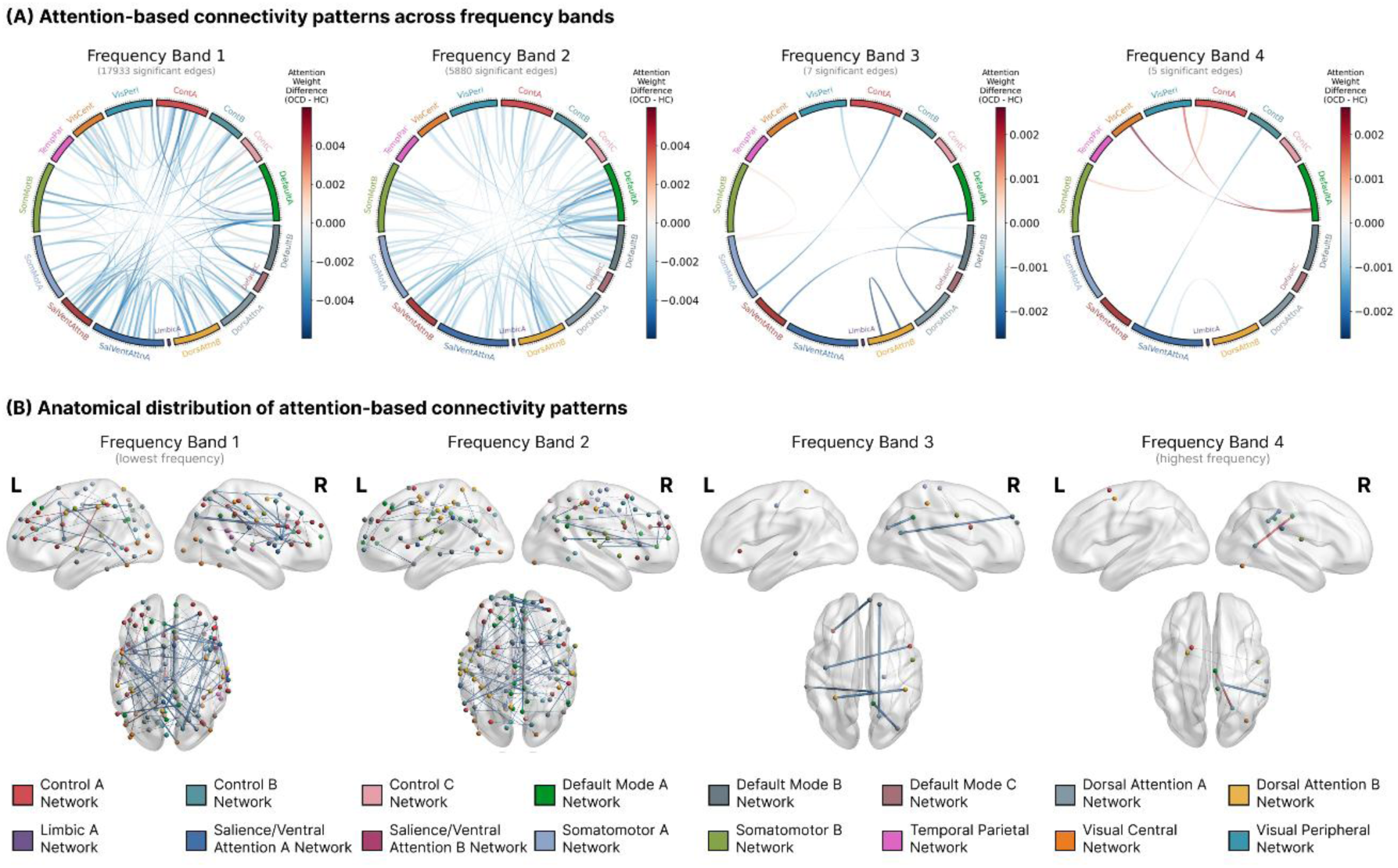
Attention-based functional connectivity (FC) differences in OCD. Significant FC edges (*p*_FDR_ < .05) between 304 ROIs from the Schaefer 400-parcel, 17-network atlas identified via attention weights. Blue edges indicate hypoconnectivity (OCD < HC); red edges indicate hyperconnectivity (OCD > HC). For Bands 1-2, we visualized the top 100 edges with the highest effect sizes (|Cohen’s *d*|). Circos plots **(A)** and 3D brain renderings with lateral and dorsal views **(B)** show widespread hypoconnectivity in Bands 1-2. A dramatic decrease in significant edges as frequency increases, indicating low-frequency patterns (< 0.10 Hz) drive classification (Band 1: 0.004-0.04 Hz, Band 2: 0.04-0.10 Hz, Band 3: 0.10-0.17 Hz, Band 4: 0.17-0.25 Hz). **Abbreviations:** OCD: obsessive-compulsive disorder, HC: healthy controls, ROI: region of interest, FDR: false discovery rate.

Notably, the training approach influenced the connectivity patterns identified by the model. The from-scratch model identified predominantly hyperconnectivity patterns (Supplementary Figure S6), whereas the pretrained model produced the widespread hypoconnectivity patterns described above.

## Discussion

By applying the MBBN model^29^ to the largest available resting-state fMRI dataset in OCD, we demonstrate that moving beyond classification accuracy reveals a more nuanced and clinically informative picture of model behavior. The model achieved modest but competitive performance (AUROC = .653), consistent with prior ENIGMA-OCD analyses^11^. Large-scale pretraining on the UK Biobank did not improve model performance, but substantially improved calibration and reduced scanner-manufacturer bias. Prediction uncertainty was driven primarily by technical rather than clinical factors, and leave-one-site-out performance varied widely (.427-.819), exposing a persistent generalization gap. Crucially, the pretrained model’s attention weights aligned with established OCD neurobiology – particularly widespread low-frequency hypoconnectivity in the default mode, salience, and somatomotor networks – whereas the from-scratch model did not. Together, these findings illustrate both the promise and the current limitations of neuroimaging-based AI for OCD and outline the methodological advances needed before such models can be considered for clinical use.

The performance of the from-scratch MBBN model (.653 ± .039) falls within the range reported in the most closely related mega-analysis of static functional connectivity in the same consortium (.567 to .673)^11^. This performance is consistent with other ENIGMA-OCD studies using different modalities, including structural MRI^18^ and white matter diffusion estimates^19^, both of which yielded low-to-moderate accuracies. Within this study, the MBBN significantly outperformed BrainLM and showed a trend toward outperforming XGBoost that did not survive FDR correction, while performance was comparable to other baseline models. These modest AUROC values contrast sharply with the high accuracies (66% to 100%) reported in single-site studies with small, homogeneous samples and standardized protocols^4^. This performance gap underscores why raw classification accuracy is insufficient, motivating the broader model assessment performed in this study.

Clinical subgroup analyses yielded only modest gains, suggesting that reducing heterogeneity by medication status or age does not resolve the limited separability of OCD and healthy controls in multi-site neuroimaging data. Medicated and unmedicated OCD were both classified slightly better from healthy controls than the full sample, but their performances were similar. This differs from prior ENIGMA-OCD functional and structural MRI studies, in which medicated patients were more accurately distinguished from healthy controls than unmedicated patients^11,18^. However, our findings are consistent with the diffusion MRI study, where classification of adult unmedicated OCD patients versus healthy controls outperformed the full-sample analysis^19^. Together, these findings suggest that the effect of medication stratification is model- and modality-dependent, rather than uniform across ENIGMA-OCD studies. We also found that the pediatric sample showed lower performance with greater variability than the adult sample, which is consistent with previous ENIGMA-OCD functional MRI study^11^ and may reflect smaller sample sizes, greater developmental heterogeneity, and the use of a transformer model, which is generally more data-hungry than conventional machine learning approaches^11,19,31^. This suggests that substantially larger and more developmentally homogeneous pediatric datasets will likely be required to reliably evaluate deep learning models in pediatric OCD.

Given these modest and variable classification performances, clinical AI models should be evaluated not only by how accurately they classify patients, but also by how reliably they express confidence in their predictions^21,22,24^. We found that pretraining substantially improved model calibration, producing fewer overconfident predictions in the 90-100% confidence range compared to the from-scratch model. This reduction in high-confidence errors is particularly important in psychiatry, where clinicians may be more likely to trust an incorrect prediction when it is accompanied by strong model confidence^22,24^. Pretraining therefore appeared to improve the model’s internal estimate of its own reliability, even when it did not improve case-control discrimination. This dissociation between discrimination and calibration suggests that large-scale pretraining may be more useful for improving the safety and trustworthiness of psychiatric AI systems than for increasing raw classification performance alone.

In the clinical environment, uncertainty-based referral could accelerate workflows by identifying the most challenging cases and referring them to human experts^32^. In other medical fields, the referral of 2-20% of the most uncertain cases significantly improved model performance and enabled it to surpass the sensitivity and specificity requirements for clinical translation^24,32^. However, our decision referral analysis found that uncertainty-based referral did not improve performance compared to random referral. This suggests that we need models with higher classification performance and better alignment between the model confidence and its actual accuracy to achieve reliable uncertainty-based referral. To this end, we must first understand what drives prediction uncertainty in our models.

Our regression analysis revealed that technical variables, including scanner manufacturer, head motion, and data quality metrics, systematically predicted higher subject-level uncertainty. Interestingly, uncertainty did not differ between OCD patients and healthy controls, suggesting that it primarily reflects variability in the imaging data, rather than ambiguity in the diagnostic signal itself. Within the patient group, medication use was associated with lower uncertainty, suggesting that predictions for medicated patients were more consistent across model iterations, even though this did not translate into better classification performance relative to unmedicated patients. Overall, these findings suggest that uncertainty in this study primarily reflects inherent data variability rather than model uncertainty that could be reduced through better training^21^.

Site-level acquisition parameters also contributed to uncertainty: sites with lower repetition times showed systematically higher uncertainty scores, possibly because these acquisition characteristics were underrepresented in the training set. Together with the strong effects of head motion and data quality, this suggests that prediction uncertainty may be reduced through stricter acquisition protocols and quality control procedures. However, in cross-sectional multi-site datasets such as ENIGMA-OCD, these sources of variability are difficult to eliminate completely. Nevertheless, identifying them provides important insight into how technical and biological heterogeneity influences model behavior, emphasizing the need for methods that explicitly account for such variability rather than assuming homogeneous data^33^.

This site-level variability poses the most significant challenge: generalizing psychiatric AI across different populations. Our leave-one-site-out validation showed a large generalization gap with performance varying from .427 to .819 across 23 sites. This demonstrates that models trained on aggregated multi-site data cannot yet reliably generalize to individual unseen sites – the exact scenario they would encounter during clinical deployment^17,20^. We hypothesized that pretraining would bridge this gap^28,33^, but the pretrained model did not significantly improve leave-one-site-out performance compared to the from-scratch model. Nevertheless, pretraining eliminated the significant scanner manufacturer bias seen in the from-scratch model, suggesting that large-scale pretraining on diverse normative data helps the model learn representations that are more invariant to specific hardware characteristics, even when it does not improve overall generalization accuracy.

Why did pretraining fail to improve generalization? One possible explanation is the substantial domain mismatch between the UK Biobank dataset used for pretraining and the heterogeneous multi-site ENIGMA-OCD dataset. UK Biobank pretraining likely captures dominant sources of variation in largely healthy, older adult brains, which may not align with OCD-relevant neural alterations. This mismatch also extends to temporal acquisition properties: although both datasets were standardized to the same input length, ENIGMA-OCD included substantial variability in scan duration and repetition time, whereas UK Biobank data were more temporally uniform. Because transformer architectures are sensitive to temporal structure, these differences may have reduced the transferability of pretrained representations^34^. Additionally, the UK Biobank dataset has relatively limited scanner and site diversity compared to ENIGMA-OCD, and pretraining on more heterogeneous, multi-site data may improve model generalization more effectively than simply increasing the number of pretraining samples^35^.

In addition to the mismatch between the pretraining and target datasets, substantial heterogeneity also exists within ENIGMA-OCD itself. According to the “different-subpopulation model”, multi-site datasets may represent several distinct subpopulations rather than noisy samples of a single population^16^. This violates the fundamental assumption that training and test samples are identically distributed, creating a domain shift problem when models generalize to unseen distributions^33^. Previous studies have addressed this using data harmonization approaches (e.g., ComBat^36^), which remove site effects as a preprocessing step to make sites appear statistically identical. However, neuroimaging ENIGMA-OCD studies found that ComBat harmonization fails to improve, and sometimes hinders, classification performance^19,37^. This failure likely stems from a fundamental limitation: data harmonization methods assume site differences are “nuisance” variables, but these differences might also contain biologically meaningful and useful information for models to learn^16,38^. We must therefore move from data harmonization to domain generalization approaches, which build models robust to site differences^33,39^. Studies on autism spectrum disorder have shown that domain generalization methods can improve model generalizability in multi-site data^40^. Building models robust to domain shifts is necessary for clinical translation, where each patient represents a domain shift problem.

Beyond generalizability, clinical AI must also ensure that models learn biologically meaningful patterns rather than spurious technical artifacts. We used frequency band decomposition to separate resting-state signals into four frequency bands, based on evidence that four bands provide the most reproducible decomposition of BOLD signals^30^ and our own control analyses. We found that the pretrained model predominantly utilized low-frequency connectivity patterns (below 0.10 Hz), yielding widespread hypoconnectivity in OCD patients that involved the default mode, salience, and somatomotor networks. The sparse high-frequency findings were observed in both from-scratch and pretrained models, suggesting that low frequencies carry more OCD-discriminative information rather than reflecting an artifact of pretraining. Altogether, these results align with network-based models implicating interactions among the default mode, salience, and executive control systems^10,41^, previous ENIGMA-OCD analyses reporting somatomotor hypoconnectivity^11^, and studies highlighting the importance of low-frequency bands (< 0.08 Hz) in OCD^15^.

However, this correspondence with prior ENIGMA-OCD findings is partial, as our model was restricted to cortical ROIs to ensure compatibility with the UK Biobank pretraining dataset. As a result, the model could not capture thalamo-cortical or other subcortical contributions, even though prior ENIGMA-OCD resting-state analyses highlighted the thalamus – particularly its hyperconnectivity with sensorimotor regions – as an important feature of OCD^11^. Training methodology also significantly influenced the identified cortical patterns: while the from-scratch model achieved the best raw accuracy, its attention maps frequently identified hyperconnectivity patterns that diverged from established neurobiological literature. In contrast, the pretrained model produced connectivity patterns more closely aligned with previous studies, suggesting that pretraining guides the model toward biologically plausible patterns. This highlights that raw performance alone is insufficient: we must know the rationale behind model’s predictions to create trustworthy AI systems.

Several methodological limitations may have influenced our results. The exclusion of sites with repetition times exceeding 2 seconds and the application of low-pass filtering at 0.25 Hz were necessary to ensure frequency band consistency but may have restricted information available to the model. Additionally, matching the UK Biobank and ENIGMA-OCD datasets required using only cortical ROIs and fixed time-series lengths, limiting the analyses of subcortical structures such as the thalamus and striatum and necessitating time series truncation or zero-padding to accommodate variable scan durations across sites. The choice of cortical brain parcellation may also have influenced both model performance and the specific connectivity patterns identified. Finally, while our attention-based connectivity analysis validates that the model uses biologically plausible signals, more rigorous interpretability methods are needed to move from model validation toward the generation of novel neurobiological hypotheses.

This study represents a critical shift toward reliable clinical AI by moving beyond simple accuracy metrics to evaluate the three pillars of trustworthy systems: reliability, interpretability, and generalizability. However, challenges remain: the current model performance is insufficient for clinical translation, the generalization gap persists leading to wide performance variability across sites, and calibration remains insufficient to integrate model uncertainty into clinical workflows. Future studies should investigate diverse pretraining strategies on large-scale heterogeneous datasets, develop methods to address data and model uncertainty, and apply domain generalization approaches to build models inherently robust to site differences. By pursuing these directions, the field can move toward AI systems that not only achieve competitive performance but also demonstrate the reliability, interpretability, and generalizability needed for safe clinical translation.

## Methods

### 1. Study participants

#### 1.1. ENIGMA-OCD cohort (primary analysis dataset)

We analyzed data from the ENIGMA-OCD Working Group, comprising 36 independent samples from 24 institutions worldwide (described in detail in Bruin et al., 2023^11^). OCD diagnoses in the data were established via structured interviews using the Diagnostic and Statistical Manual of Mental Disorders-IV (DSM-IV)^2^ or DSM-5^3^ (see Supplementary Methods for details). Symptom severity was assessed using the Yale-Brown obsessive-compulsive scale (Y-BOCS)^42^ and the Children’s Y-BOCS^43^.

The initial dataset included 3,044 subjects. We excluded 99 subjects without neuroimaging data, 36 without diagnosis, sex, or age information, two healthy controls (HCs) using medication at scan time, and 12 HCs with positive Y-BOCS values. After this covariate quality control (N = 2,895), we excluded 268 subjects who failed neuroimaging quality control, then 115 due to excessive motion, 244 with insufficient brain coverage or missing time-series values, and 174 from samples with fewer than 10 subjects per group across two quality control stages. We also excluded 388 subjects from 6 samples with repetition time (TR) > 2 seconds, as long TRs prevent obtaining the full frequency range (0.01–0.25 Hz) required for accurate frequency band decomposition^30^ (see Supplementary Figure S1 for study flowchart).

The final dataset included 1,706 participants (869 OCD patients, 837 HCs). Demographic and clinical characteristics of this dataset are summarized in Table 1, and details on site-specific characteristics, comorbidities, and symptom dimensions are in Supplementary Tables S1-S2.

#### 1.2 UK Biobank cohort (pretraining dataset)

For the pretraining, we used resting-state functional magnetic resonance imaging (rs-fMRI) data from the UK Biobank (UKB) cohort^44^. Among 41,283 participants, we excluded 491 participants with zero variance in at least one region of interest (ROI) and 9 participants whose data contained missing values after frequency band decomposition. The final pretraining dataset included 40,783 participants (mean age: 55.0 ± 7.5 years, range: 40–70 years; 47.0% male).

### 2. Data acquisition and preprocessing

#### 2.1 Image acquisition and preprocessing

ENIGMA-OCD rs-fMRI images were acquired using Siemens, Philips, or GE scanners of the 1.5 or 3 Tesla field strength. Initial TRs ranged from 0.7 to 3.5 seconds and scan durations from 4 to 12 minutes. The final dataset had TRs from 0.7 to 2.0 seconds and voxel resolutions of 2-3.5 mm isotropic after excluding sites with TR > 2 seconds (Supplementary Table S3). Images were preprocessed using the Harmonized AnaLysis of Functional MRI pipeline (HALFpipe)^45^, versions 1.0.0-1.2.1, following standardized ENIGMA protocols: motion correction, slice-timing correction, susceptibility distortion correction (where applicable), and spatial normalization to the Montreal Neurological Institute template (MNI152). Denoising included motion artifact removal via independent component analysis-based automatic removal of motion artifacts (ICA-AROMA)^46^, spatial smoothing (6 mm full-width at half-maximum Gaussian kernel), and regression of the top five anatomical component correction (aCompCor) components^47^. Acquisition and preprocessing details are described in Bruin et al. (2023)^11^.

UKB rs-fMRI images were acquired on Siemens Skyra 3T scanners (TR = 0.735 s, voxel size = 2.4×2.4×2.4 mm, duration = 6 minutes, 490 volumes) and preprocessed using the standard UKB pipeline^48^: motion correction, grand-mean intensity normalization, high-pass temporal filtering, echo-planar imaging unwarping, structured artifact removal, and spatial normalization to MNI space.

#### 2.2 Frequency filtering

For the ENIGMA-OCD dataset, we applied a low-pass Butterworth filter (4th order, zero-phase, cutoff = 0.25 Hz) to standardize the maximum frequency across all subjects. Filtering was a necessary step; otherwise, TRs ranging from 0.7 to 2.0 seconds across sites would result in Nyquist frequencies from 0.25 to 0.71 Hz, producing inconsistent frequency boundaries across subjects during band decomposition (Section 3). As a control analysis, we also trained the model on unfiltered data (Supplementary Tables S5). UKB data had undergone bandpass filtering from 0.008 to 0.1 Hz during preprocessing. Notably, this means that the UKB pretraining data contained no signal above 0.1 Hz, creating a frequency mismatch with Bands 3 and 4 of the ENIGMA-OCD decomposition. As a control analysis addressing this mismatch, we trained both from-scratch and pretrained models on ENIGMA-OCD data filtered to the 0.01-0.1 Hz range, which showed classification performance slightly lower than main analyses (from-scratch AUROC = .625 ± .039; pretrained AUROC = .622 ± .045, across three random seeds; see Supplementary Table S4).

#### 2.3 Brain parcellation and time series preprocessing

ENIGMA-OCD data used a multi-atlas parcellation with a total of 434 ROIs: 400 cortical ROIs from the Schaefer atlas^49^, 17 subcortical ROIs from the Harvard-Oxford atlas^50^, and 17 cerebellar ROIs from the Buckner atlas^51^. After quality control for insufficient echo-planar imaging coverage and missing values, 318 ROIs remained (see details in Supplementary Methods).

UKB data initially included 400 cortical ROIs from the Schaefer atlas^49^. To enable direct transfer learning between UKB and ENIGMA-OCD, we constrained the feature space to the cortical Schaefer parcellation available in both datasets. This design prioritized cross-dataset harmonization and stable transfer learning, but led to the exclusion of subcortical and cerebellar ROIs, including those reported relevant to OCD in previous studies^11,12^.

We further removed three ROIs with the lowest blood-oxygen-level-dependent (BOLD) signal variance to satisfy architectural requirements of the MBBN model (i.e., the number of ROIs must be divisible by the number of attention heads)^29^. This procedure yielded the final set of 304 cortical ROIs. Mean BOLD time series were extracted for each ROI by averaging voxel signals within the ROI at each time point.

For the temporal dimension, we removed the first 20 volumes from both datasets for T1 equilibration. To ensure consistency between datasets and satisfy architectural constraints of the MBBN model (i.e., time series length must be divisible by the number of attention heads)^29^, we standardized the target time-series length to 464 time points. ENIGMA-OCD time series were matched to this length: longer time series were truncated, and shorter ones were zero-padded. Subjects with missing values in the time-series data were excluded.

### 3. Frequency band division strategy

The original MBBN framework decomposes time-series data into three frequency bands using Lorentzian and spline multifractal fitting^29^. In contrast, we used variational mode decomposition (VMD)^52^ to separate resting-state fMRI (rs-fMRI) signals into four frequency bands, known as intrinsic mode functions. Each function is defined by a specific central frequency and bandwidth. We chose VMD because the original MBBN strategy was designed for signals bandpass filtered in ∼0.01-0.1 Hz range and yielded inconsistent band definitions on the ENIGMA-OCD data with a higher Nyquist frequency (0.25 Hz). VMD offers a more robust alternative for rs-fMRI data across varying frequency ranges^30^.

For each participant, we applied VMD to the mean time series across all ROIs to determine frequency band boundaries. We then used these boundaries to extract frequency bands from each ROI’s time series via bandpass filtering. We set the number of bands to four based on a previous study where four bands produced the most precise and reproducible decomposition of rs-fMRI signals^30^. Our control analyses further confirmed this choice, comparing configurations with one, two, three, and five frequency bands and yielding the best classification performance with four frequency bands (Supplementary Table S6). We optimized the VMD parameters for the ENIGMA-OCD and UKB datasets to minimize reconstruction error, following the original VMD approach^52^ (Supplementary Table S7). This procedure generated four distinct time series per brain region, which served as the primary input for the model (see Supplementary Table S8 for band cutoffs).

### 4. Deep learning model architecture (Multi-Band Brain Net)

We adapted the Multi-Band Brain Net (MBBN), a transformer-based architecture that models frequency-specific spatiotemporal brain dynamics^29^. MBBN processes time series from 304 brain regions for each participant, with each region’s time series decomposed into four frequency bands (Figure 1). The model consists of two modules operating within each band.

The temporal module captures temporal dynamics using a bidirectional encoder representations from transformers (BERT) encoder^53^ with the same neural network weights shared across all bands. This module produces logits (*y*^_*j*_) for each frequency band *j*. The final classification prediction (*y*^) is obtained by averaging the logits from each frequency band:

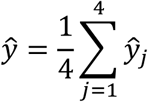

The spatial module learns frequency-specific patterns of dynamic functional connectivity via multi-head attention mechanisms and uses separate neural network weights for each frequency band^29^. This design choice - shared weights in the temporal module and independent weights in the spatial module - allows the model to learn both common temporal dynamics and frequency-specific spatial connectivity patterns. Attention matrices from the spatial module further reveal which functional connections are most important for classification at each frequency band.

The temporal and spatial modules operate in parallel on the input data and are coupled through a composite loss function. This loss function combines a classification-task-specific loss with a spatial loss^29^:

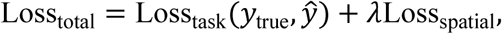

where *y*_true_ represents the true diagnostic labels (OCD or healthy control), *y*^ represents the model’s predicted probability of OCD, and spatial loss factor, *λ*, controls how much weight is given to the spatial loss relative to the task loss. The task loss (Loss_task_) is a binary cross-entropy loss that measures how well the model’s predictions match the true labels. The spatial loss (Loss_spatial_) encourages the model to learn unique patterns of brain connectivity for each frequency band.

### 5. Model training and evaluation

We investigated two strategies to train the MBBN model for OCD classification. First is training the model from scratch on the ENIGMA-OCD dataset with randomly initialized weights. Second is pretraining the model on the UKB dataset first and then finetuning it on the ENIGMA-OCD dataset.

#### 5.1 Pretraining procedure (UK Biobank)

To examine if large-scale pretraining enhances model generalizability, we pretrained the MBBN on the UKB resting-state fMRI dataset (N = 40,783; Section 1.2) using a self-supervised task called masked signal modeling^29^. This task trains the model to learn fundamental brain dynamics by reconstructing masked segments of the fMRI signal. We implemented a dual-masking strategy: spatial masking targeted high-communicability hub regions across all four frequency bands, while temporal masking removed consecutive time windows. By reconstructing these missing segments from the surrounding context, the model learns to capture complex spatiotemporal dependencies inherent in neural activity.

We generated 64 pretrained model candidates by systematically varying three masking parameters: the number of masked ROIs, the width of the temporal masking windows, and the spacing between masked windows. We then conducted a two-tier selection process to identify the optimal pretrained model configuration for finetuning. Complete pretraining procedures and optimization details are provided in the Supplementary Methods.

#### 5.2 OCD classification and model evaluation (ENIGMA-OCD)

We trained from-scratch and pretrained MBBN models on the ENIGMA-OCD dataset to classify OCD patients from healthy controls. We randomly split the dataset into training, validation, and test sets in a 70:15:15 ratio. The training set was used to train the model, the validation set was used for hyperparameter optimization, and the test set was reserved for final performance evaluation. To prevent class imbalance from influencing model training or evaluation, we stratified the splits by OCD diagnosis. No subjects overlapped between the train, validation, and test sets. Classification performance was evaluated using the area under the receiver operating characteristic curve (AUROC) and balanced accuracy.

To ensure robustness, we repeated the data splitting procedure 10 times using different random seeds. Hyperparameters were optimized using an automated search procedure (Optuna^54^) during model development. The validation set from the first split was used to determine the optimal hyperparameters before observing test performance, and these hyperparameters were used in all subsequent experiments to ensure consistent evaluation across splits. Complete hyperparameter optimization procedures and final hyperparameter settings are provided in Supplementary Table S9.

We performed statistical analyses to check: (1) whether model performance significantly exceeded chance-level classification, and (2) whether there were significant performance differences between models. Because the 10 AUROC values per model were obtained from overlapping random splits, standard tests assuming independent observations would underestimate variance and inflate Type I error. We therefore applied the Nadeau-Bengio corrected paired t-test^55^, which adjusts the variance estimate by the factor 1/*n* + *ρ*, where *n* = 10 is the number of splits and *ρ* = *n*_*test*_/*n*_*train*_ reflects the ratio of test to training set size. We used a one-sided Nadeau-Bengio t-test to assess whether model performance exceeded chance and a two-sided test with false discovery rate (FDR) correction for pairwise model comparisons. The number of splits was constrained by the high computational cost of training deep learning models, but we acknowledge that 10 splits may provide limited statistical power to detect small to moderate effect sizes.

To evaluate model robustness across clinically relevant subgroups, we performed additional classification analyses using 10 random splits for each analysis. These subgroup analyses assessed whether the model maintained stable performance across different patient populations that may exhibit distinct neural signatures. Specifically, we evaluated performance for medicated patients (447 OCD vs 837 HC), unmedicated patients (415 OCD vs 837 HC), adult sample (760 OCD vs 754 HC), and pediatric populations (109 OCD vs 83 HC). This approach allowed us to determine whether the model’s classification performance generalized across diverse clinical presentations and developmental stages.

### 6. Benchmarking and baseline comparisons

We compared MBBN against several baseline models using the same 10 data splits (Section 5.2) to ensure fair comparison. We selected these baselines to test two primary hypotheses: first, whether direct processing of raw time-series data outperforms traditional static functional connectivity (FC) features; and second, whether our multi-band decomposition provides a significant advantage over existing single-band or foundation-model architectures.

The first group of baseline models utilized pre-computed static FC matrices. These matrices were computed by calculating Pearson correlation coefficients between all pairs of ROI time series, followed by Fisher’s z-transformation. This category included traditional machine learning benchmarks – extreme gradient boosting (XGBoost)^56^ and support vector machine (SVM)^57^ – together with the more recent brain network transformer (BNT)^58^, which applies attention mechanisms to connectivity graphs.

The second group of baseline models directly processed raw time-series fMRI data. It included blood-oxygen-level-dependent transformer (BoIT)^59^, vanilla BERT^53^ (a standard BERT encoder without MBBN’s multi-band structure or spatial module), and BrainLM^60^ (a foundation model pretrained on large-scale fMRI data and finetuned on ENIGMA-OCD data).

BrainLM was originally pretrained on datasets with highly similar temporal resolutions and does not explicitly model variable TRs^60^. In contrast, the ENIGMA-OCD dataset included substantial TR heterogeneity across sites. To ensure comparability across models, we provided identical preprocessed inputs to all models that use raw time-series data, rather than applying BrainLM-specific temporal resampling. Therefore, BrainLM’s relatively lower performance may partly reflect a mismatch between its pretraining conditions and the heterogeneous characteristics of the ENIGMA-OCD dataset.

Detailed hyperparameter optimization procedures and final model configurations are provided in Supplementary Methods and Supplementary Table S10, respectively.

### 7. Uncertainty quantification

Understanding when a model is uncertain about its predictions is essential for the safe translation of deep learning into clinical psychiatry. Identifying cases where predictions are unreliable helps determine when a model requires expert human evaluation or alternative diagnostic tools^21,24,61^.

While current performance levels across the field do not yet support direct clinical use, we implemented uncertainty quantification to evaluate prediction reliability and identify factors influencing model confidence. This approach provides a framework for developing more trustworthy, transparent and cautious tools for future clinical decision support.

#### 7.1 Estimating prediction uncertainty

We estimated prediction uncertainty using Monte Carlo dropout (MC dropout) – a widely used uncertainty quantification method for deep learning models^21,22^. This method approximates Bayesian inference by enabling dropout layers during test-time. For each test subject, we performed 100 forward passes through the trained network with dropout layers kept active during inference, using the same dropout probability as during training. The final prediction probability was the average across all 100 passes, and we quantified prediction uncertainty as the variance of these predictions. This approach allows the model to capture predictive uncertainty when outputs vary substantially across passes, avoiding the single overconfident predictions common in traditional architectures^21,24^.

#### 7.2 Assessing model calibration

Model calibration measures the alignment between a model’s confidence and its actual accuracy. For instance, when the model predicts 80% confidence, does it achieve 80% accuracy? Poorly calibrated models often display over- or underconfidence, which can mislead clinicians into either over-relying on incorrect model predictions or dismissing accurate predictions^21,23,24^.

We assessed calibration using calibration plots (“reliability diagrams”) to visualize how well predicted confidence matches empirical accuracy. We grouped predictions into 10 bins based on confidence scores (e.g., 0–10%, 10–20%) and calculated the average predicted confidence and actual accuracy for each bin.

We quantified calibration using two metrics: expected calibration error (ECE), the weighted average gap between confidence and accuracy across all bins, and maximum calibration error (MCE), the largest gap in any single bin. Lower ECE and MCE values indicate better calibration. We compared calibration between pretrained and from-scratch MBBN models using paired Wilcoxon signed-rank tests for both ECE and MCE. Unlike the classification comparisons (Section 5.2), we retained the Wilcoxon signed-rank test rather than applying the Nadeau-Bengio correction, given the larger number of splits and the magnitude of the observed effects.

#### 7.3 Identifying predictors of model uncertainty

To identify factors influencing model uncertainty, we used multiple linear regression predicting log-transformed uncertainty scores. We included a comprehensive range of potential influences such as clinical predictors (OCD diagnosis, medication status, symptom severity, and comorbidities), demographic variables (age and sex), and technical predictors (scanner type, head motion and data quality metrics). We fitted separate models for the whole sample, healthy controls, and OCD patients. Continuous predictors were standardized before model fitting. We set statistical significance at *p* < .05 and applied corrections for multiple comparisons where appropriate.

#### 7.4 Post-hoc analyses

We performed three post-hoc analyses to characterize uncertainty patterns in model predictions and evaluate whether uncertainty could improve clinical decision-making. First, we compared uncertainty distributions between pretrained and from-scratch models using Wilcoxon signed-rank tests. Second, we evaluated whether uncertainty differed between healthy controls and OCD patients across clinical subgroups (e.g., symptom dimensions, comorbidity status) to determine if the model was more uncertain about specific phenotypes. Third, we assessed whether referring high-uncertainty cases for expert review improved classification performance compared to random exclusion, simulating a clinical workflow where uncertainty informs human intervention. Specifically, we systematically excluded predictions with the highest uncertainty and recalculated performance metrics on the remaining, more confident samples. This analysis quantifies the gain in diagnostic reliability achieved when the model is permitted to defer its most ambiguous cases to clinical experts^24,32^.

### 8. Model generalizability

#### 8.1 Leave-one-site-out cross-validation procedure

We assessed model generalizability using leave-one-site-out cross-validation (LOSO-CV) across 23 sites in the ENIGMA-OCD dataset. In each fold, we held out one site as the test set and used the remaining 22 sites for training and validation. This approach evaluates the model on completely unseen data, testing generalizability across different scanners, acquisition parameters, and site-specific protocols. To ensure robustness, we repeated each fold three times with random subject shuffling between training and validation sets, yielding three performance estimates per site.

We performed statistical comparisons to determine if pretraining enhanced generalizability and if MBBN models outperformed established baselines. Specifically, we compared median LOSO performance between from-scratch and pretrained MBBN models using paired Wilcoxon signed-rank tests. We evaluated performance across all models using Friedman tests followed by pairwise post-hoc comparisons with multiple comparisons correction. All tests used a significance level of *p* < .05.

#### 8.2 Quantifying site-level heterogeneity in functional connectivity

We quantified site-level heterogeneity in the fMRI data using intra-class correlation (ICC) analysis. This analysis measures the proportion of variance in functional connectivity attributable to site-level factors – such as scanner characteristics and acquisition parameters – versus within-site subject-level differences.

For each of the 1,706 subjects, we computed functional connectivity between all 304 pairs of brain regions, yielding 46,056 unique connections per subject. We normalized these values using Fisher’s z-transformation. For each connection, we decomposed the total variance into site-specific and residual (individual) components to calculate the ICC:

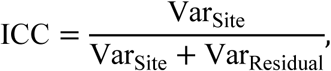

where Var_Site_ represents variance across sites and Var_Residual_ captures individual differences and measurement error. This procedure yields an ICC value for each of the 46,056 connections. The values range from 0 to 1, where higher scores indicate that site differences explain a larger proportion of the variance in connectivity patterns.

#### 8.3 Scanner manufacturer effects on model generalizability

Scanner manufacturer variations introduce substantial domain shift in multi-site neuroimaging studies, with deep learning models often achieving high accuracy at identifying scanner manufacturers and showing a significant performance drop when generalizing across different hardware^17^. This manufacturer-specific bias poses a critical barrier to clinical translation, where models must generalize across diverse scanner environments. To evaluate whether pretraining mitigates this technical confound, we assessed whether LOSO-CV performance varied across scanner manufacturers (GE, Siemens, or Philips) for both the from-scratch and pretrained MBBN models. We performed separate one-way analysis of variance (ANOVA) tests on site-level AUROC scores for each model, grouping sites by scanner manufacturer. This analysis allowed us to determine whether pretraining reduces manufacturer-specific bias by learning features more invariant to scanner hardware.

#### 8.4 Uncertainty patterns across sites

We analyzed uncertainty estimates across sites to understand how technical acquisition parameters influence model confidence. For each site, we averaged the Monte Carlo dropout-based uncertainty scores (Section 7.1) across all subjects from that site to obtain site-level uncertainty estimates. We then examined whether technical acquisition factors such as repetition time (TR) or scanner manufacturer were associated with site-level model uncertainty. To assess this, we visualized site-level uncertainty together with LOSO-CV performance across sites to explore whether acquisition parameters were associated with systematic differences in model confidence.

### 9. Model interpretability

#### 9.1 Attention-based functional connectivity patterns

To identify the brain connectivity patterns driving classification, we analyzed the attention weights from the spatial module. This module learns functional connectivity matrices that represent the importance of interactions between brain regions for distinguishing OCD patients from healthy controls (HCs). These learned matrices reveal which brain region pairs are most important for classification at each frequency band.

#### 9.2 FC extraction and statistical comparison

To ensure the interpretability of our results, we focused on subjects correctly classified across all random seeds where they appeared in a test set (113 controls and 254 cases with OCD for the from-scratch model; 139 controls and 110 cases with OCD for the pretrained model). For these individuals, we extracted the 304 × 304 attention matrices for each of the four frequency bands, averaging weights across attention heads and random seeds to obtain a single functional connectivity matrix per subject per band.

We compared these attention-based patterns between OCD and HC groups using edge-wise Mann-Whitney U tests for each frequency band. We calculated effect sizes using Cohen’s *d* and applied multiple comparisons correction to edges from each frequency band. We retained edges with an adjusted *p* < .05 that showed significant differences in either direction. Finally, we visualized these significant connectivity alterations using circular plots and three-dimensional brain renderings^62^. We used these results to validate that the models learned biologically plausible features by comparing the significant connectivity alterations to established literature on functional network alterations in OCD.

### 10. Ethical approval

This study utilized anonymized data from the ENIGMA-OCD consortium and the UK Biobank. All procedures contributing to this work comply with the ethical standards of the relevant national and institutional committees on human experimentation and with the Helsinki Declaration of 1975, as revised in 2008. Ethical approval and informed consent were obtained by the original data collection initiatives at each participating site, as detailed in the respective cohort publications and participant information sections above.

### 11. Data availability

The ENIGMA-OCD consortium data are available to researchers upon reasonable request and completion of a data use agreement (contact details available via the ENIGMA consortium website: http://enigma.ini.usc.edu/). UK Biobank data are accessible to registered researchers via application through the UK Biobank Access Management System (https://www.ukbiobank.ac.uk/).

### 12. Code availability

The code used for model training, evaluation, and all post-hoc analyses is available at https://github.com/Transconnectome/enigma_ocd_mbbn.git.

## Acknowledgments

This work was supported by the National Research Foundation of Korea (NRF) grant funded by the Korea government (MSIT) (No. 2021R1C1C1006503, RS-2023-00266787, RS-2023-00265406, RS-2024-00421268, RS-2024-00342301, RS-2024-00435727, RS-2025-25457239, RS-2021-NR061370, NRF-2021M3E5D2A01022515, and NRF-2021S1A3A2A02090597), by the Researchers Program through Seoul National University (No. 200-20250071, 200-20250049, 200-20250116, 200-20250115, 200-20250113, 0670-20250039, 200-20260009). Additional support was provided by the Institute of Information & Communications Technology Planning & Evaluation (IITP) grant funded by the Korea government (MSIT) [No. RS-2021-II211343, Artificial Intelligence Graduate School Program, Seoul National University] and by the Global Research Support Program in the Digital Field (RS-2024-00421268). This work was also supported by the Artificial Intelligence Industrial Convergence Cluster Development Project funded by the Ministry of Science and ICT and Gwangju Metropolitan City, by the Korea Brain Research Institute (KBRI) basic research program (25-BR-05-01), by the Korea Health Industry Development Institute (KHIDI) and the Ministry of Health and Welfare, Republic of Korea (HR22C1605), and by the Korea Basic Science Institute (National Research Facilities and Equipment Center) grant funded by the Ministry of Education (RS-2024-00435727). We acknowledge the National Supercomputing Center for providing supercomputing resources and technical support (KSC-2023-CRE-0568, KSC-2024-CRE-0198, KSC-2025-CRE-0340). An award for computer time was provided by the U.S. Department of Energy’s (DOE) ASCR Leadership Computing Challenge (ALCC). This research used resources of the National Energy Research Scientific Computing Center (NERSC), a DOE Office of Science User Facility, under ALCC award m4750-2024, and supporting resources at the Argonne and Oak Ridge Leadership Computing Facilities, U.S. DOE Office of Science user facilities at Argonne National Laboratory and Oak Ridge National Laboratory. Additional funding was supported by AMED Brain/MINDS Beyond Program (Grant No. JP18dm0307002) and JSPS KAKENHI Grants No. JP19K03309, JP22H01090, JP23K07004, JP23K22361, JP23K02956, JP24K21493, JP25H01085, JP25K00879 awarded to YH; Sao Paulo Research Foundation (FAPESP Grants No. 25/19467-3, 23/18337-3, 24/09675-5, 23/16997-6, 21/05332-8 and 23/02538-0) to JRS; the Marató TV3 Foundation Grant No. 202201-30 and Carlos III Institute of Health Grants No. PI22_00752 and PI25_01407 to PA; Grant No. RYC2021-031228-I funded by Spanish Ministerio de Ciencia e Innovación (MCIN/AEI/10.13039/501100011033) and by the European Union NextGenerationEU/PRTR to MPP; Japan Society for the Promotion of Science (KAKENHI Grant No. 23K14825) to YA; AMED under Grant No. JP23wm0625001 and JP24wm0625204 to YS; Wellcome-DBT India Alliance Grant No. 500236/Z/11/Z to GV; South African Medical Research Council (MRC); a grant from the SA National Research Foundation (NRF) to CL; Grants No. R01MH126981, R01MH111794, and R33MH107589 from the National Institute of Mental Health/National Institute of Health to ERS; NIH Grant No. R01MH138569 to PMT and SIT; and LW was supported by German Center for Mental Health (DZPG).

