## Supplementary Material for "Toward trustworthy clinical AI for obsessive-compulsive disorder: reliability, generalizability, and interpretability of a transformer model across the ENIGMA-OCD consortium"

**This file includes:**

Supplementary Methods
References
Supplementary Figures S1-S6
Supplementary Tables S1-S13

### Supplementary Methods

#### 1. Diagnosis instruments

The diagnosis of OCD was established according to DSM-IV or DSM-5 criteria using structured interviews (adult samples: Mini-International Neuropsychiatric Interview (MINI)[1] and Structured Clinical Interview for DSM Disorders (SCID)[2]; pediatric samples: Schedule for Affective Disorders and Schizophrenia for School-Age Children – Present and Lifetime Version (K-SADS-PL)[3], MINI for Children and Adolescents (MINI-KIDS)[4], and Anxiety Disorders Interview Schedule (ADIS)[5]).

#### 2. Neuroimaging data quality control (QC)

Following covariate QC (N = 2,895), rigorous magnetic resonance imaging (MRI) QC was applied based on standardized ENIGMA functional protocols (http://enigma.ini.usc.edu/protocols/functional-protocols/). This led to the exclusion of 268 subjects due to issues with T1w skull stripping, spatial normalization, echo planar imaging (EPI) signal-to-noise ratio, confound time series, or independent component analysis (ICA) noise components. An additional 115 subjects were excluded based on motion criteria: any rotation/translation parameter exceeding 4 mm/degrees, average framewise displacement (FD) > 0.3 mm, or fewer than 100 volumes of motion-unaffected data (FD < 0.25 mm). Subsequently, 79 subjects were removed from 4 samples with fewer than 10 examples per class.

Further QC was performed on the remaining 2,433 participants and an initial set of 434 regions of interest (ROIs). First, two ROIs were excluded due to zero EPI coverage across participants, leaving 432 ROIs. Subsequently, two participants were removed as they had insufficient coverage (less than 50% voxel coverage in less than 90% of the remaining ROIs). From the data of the remaining 2,431 participants, an additional 29 ROIs were excluded because less than 90% of participants had at least 50% coverage for these regions. Furthermore, 85 ROIs were removed due to having more than 1% of participants with missing time-series values. This resulted in a refined set of 318 ROIs. Using this set of 318 ROIs, 242 participants were excluded due to the presence of any missing values in their ROI time-series data. Finally, after these ROI-based subject exclusions, an additional 95 subjects were removed because they belonged to sites with fewer than 10 participants per diagnostic group.

After neuroimaging QC, we excluded 388 subjects from 6 samples with a repetition time (TR) > 2 seconds, as these longer sampling rates preclude the frequency band decomposition (up to 0.25 Hz) required for our analysis. This yielded a dataset of 1,706 participants and 318 ROIs. To harmonize with the pretraining dataset (UK Biobank), which utilized Schaefer atlas ROIs[6], 11 non-Schaefer (subcortical and cerebellar) ROIs were removed from the ENIGMA-OCD set. To meet the MBBN model's architectural requirement that the number of ROIs be divisible by the number of attention heads (four), three additional Schaefer ROIs with the lowest BOLD signal variance across subjects were subsequently removed. Thus, the final analyses for both the ENIGMA-OCD and UKB datasets were conducted using time series from 304 common Schaefer ROIs. A full study flowchart is shown in Supplementary Figure S1.

#### 3. Frequency band division strategy

Variational Mode Decomposition (VMD)[7] was employed to decompose the BOLD time series from each of the 304 ROIs into distinct frequency-specific components. This adaptive, data-driven approach was chosen because the original Multi-Band Brain Net (MBBN) model's frequency-division strategy[8] was developed for datasets typically band-pass filtered in the ~0.01-0.1 Hz range. This strategy yielded inconsistent band definitions on the ENIGMA-OCD dataset, which is characterized by highly variable repetition times (TRs) ranging from 0.7 to 2.0 seconds. In contrast, VMD has been shown to be robust in decomposing rs-fMRI signals across different TRs[9]. The VMD was implemented using the Python package sktime version 0.35.1[10]).

##### 3.1. VMD core principles

VMD is an algorithm that decomposes a real-valued input signal, $f(t)$, into a discrete number of sub-signals, or modes, $u_k(t)$, where $k = 1, ..., K$ is the predetermined number of modes. Each mode $u_k(t)$ is called an Intrinsic Mode Function (IMF) and is mostly compact around a central frequency $\omega_k$, which is estimated concurrently with the modes themselves. An IMF is typically an amplitude-modulated-frequency-modulated (AM-FM) signal of the form:

$$u_k(t) = A_k(t)\cos(\phi_k(t))(\text{Eq. S1})$$

where $A_k(t)$ is the instantaneous amplitude (envelope) and $\phi_k(t)$ is the instantaneous phase. The instantaneous frequency is $\omega_k(t) = \frac{d\phi_k(t)}{dt}$.

The primary goal of VMD is to ensure that the sum of the extracted modes reconstructs the original signal:

$$\sum_{k=1}^{K} u_k(t) = f(t) \quad (\text{Eq. S2})$$

To achieve this, VMD seeks to minimize the sum of the bandwidths of all modes, subject to the constraint that the modes sum to the original signal. The bandwidth of each mode $u_k(t)$ is assessed by:

1. Computing its associated analytic signal via the Hilbert transform to obtain a unilateral frequency spectrum.
2. Shifting the mode's frequency spectrum to its baseband by mixing with a complex exponential tuned to its estimated center frequency $\omega_k$.
3. Estimating the bandwidth as the squared $L^2$-norm of the gradient of this demodulated signal (i.e., $H^1$ Gaussian smoothness).

The constrained variational problem is formulated as:

$$\min_{\{u_k\},\{\omega_k\}} \left\{ \sum_{k=1}^{K} \left\| \partial_t \left[ \left( \delta(t) + \frac{j}{\pi t} \right) * u_k(t) \right] e^{-j\omega_k t} \right\|_2^2 \right\} \quad (\text{Eq. S3})$$

subject to $\sum_{k=1}^{K} u_k = f$.

Here, $\{u_k\} := \{u_1, ..., u_K\}$ represents the set of all modes, and $\{\omega_k\} := \{\omega_1, ..., \omega_K\}$ is the set of their respective center frequencies. $\delta(t)$ is the Dirac delta function, and $*$ denotes convolution.

##### 3.2. VMD optimization algorithm

The VMD optimization algorithm can be summarized as follows (based on Algorithm 2 in Dragomiretskiy & Zosso, 2014[7]):

1. **Initialization:**
   a. Initialize the estimates for the modes in the Fourier domain, $\{\hat{u}_k^1\}$.
   b. Initialize the estimates for the center frequencies, $\{\omega_k^1\}$.
   c. Initialize the estimate for the Lagrangian multiplier in the Fourier domain, $\hat{\lambda}^1$.
   d. Set an iteration counter $n \leftarrow 0$.
2. **Iteration loop:** Repeat the following steps until a convergence criterion is met:
   a. **Increment the iteration counter:** $n \leftarrow n + 1$.
   b. **Update modes:** For each mode $k$ from $1$ to $K$, update the estimate of the mode $\hat{u}_k^{n+1}(\omega)$ for all

non-negative frequencies $\omega \geq 0$. This update effectively applies a Wiener filter to the residual signal (the original signal minus all other modes and adjusted by the Lagrangian multiplier), where the filter is tuned to the current center frequency $\omega_k^n$ of that mode:

$$\hat{u}_k^{n+1}(\omega) \leftarrow \frac{\hat{f}(\omega) - \sum_{i<k} \hat{u}_i^{n+1}(\omega) - \sum_{i>k} \hat{u}_i^n(\omega) + \frac{\hat{\lambda}^n(\omega)}{2}}{1 + 2\alpha(\omega - \omega_k^n)^2} \text{(Eq. S4)}$$

where $\alpha$ is a balancing parameter for data-fidelity.

c. **Update center frequencies:** For each mode $k$ from $1$ to $K$, update the estimate of the center frequency $\omega_k^{n+1}$ by calculating the center of gravity of the power spectrum of the newly updated mode $\hat{u}_k^{n+1}(\omega)$:

$$\omega_k^{n+1} \leftarrow \frac{\int_0^\infty \omega |\hat{u}_k^{n+1}(\omega)|^2 d\omega}{\int_0^\infty |\hat{u}_k^{n+1}(\omega)|^2 d\omega} \text{(Eq. S5)}$$

d. **Update lagrangian multiplier (dual ascent):** Update the Lagrangian multiplier $\hat{\lambda}^{n+1}(\omega)$ for all non-negative frequencies $\omega \geq 0$. This step enforces the reconstruction constraint:

$$\hat{\lambda}^{n+1}(\omega) \leftarrow \hat{\lambda}^n(\omega) + \tau \left( \hat{f}(\omega) - \sum_k \hat{u}_k^{n+1}(\omega) \right) \text{(Eq. S6)}$$

where $\tau$ is an update parameter that influences the strictness of the constraint enforcement.

3. **Convergence check:** The loop continues until the sum of squared differences between successive mode estimates falls below a predefined tolerance $\epsilon$:

$$\sum_k \frac{\|\hat{u}_k^{n+1} - \hat{u}_k^n\|_2^2}{\|\hat{u}_k^n\|_2^2} < \epsilon \text{(Eq. S7)}$$

##### 3.3. VMD implementation pipeline for rs-fMRI data

Our VMD pipeline was implemented as follows for each participant:

1. **Mean time series computation:** The BOLD time series from all 304 ROIs were averaged to create a single mean time series for that participant.
2. **VMD application for cutoff determination:** VMD was applied to this mean time series to determine subject-specific frequency band cutoffs.
3. **Application of cutoffs to ROI time series:** The frequency band cutoffs derived from the mean time series were then used to filter the original time series for each of the 304 ROIs. This approach ensured that frequency band definitions were consistent across all ROIs within a given participant but could vary between participants to adapt to individual signal characteristics. Applying VMD separately to each ROI to calculate unique cutoffs for each ROI for each participant was deemed computationally prohibitive for this large-scale analysis.

##### 3.4. VMD parameter settings

The VMD algorithm requires several parameters to be set.

Fixed parameters:

- $K = 4$: The number of modes (IMFs) to be extracted. This was chosen based on prior literature suggesting its optimality for rs-fMRI data[9] and confirmed by our control analyses (Supplementary Table S6).

- DC component (DC) = $0$: No specific mode was constrained to capture a DC component (i.e., $\omega_1 = 0$ was not enforced for the first mode).
- Initialization of omegas (init) = $0$: The initial center frequencies $\{\omega_k\}$ were uniformly distributed across the available frequency spectrum.
- Convergence tolerance (tol) = $1 \times 10^{-7}$: The stopping criterion for the VMD iteration.

Tuned parameters ($\alpha$ and $\tau$):

The balancing parameter $\alpha$ (bandwidth constraint) and the update parameter $\tau$ (Lagrangian multiplier update rate) were tuned for each dataset (ENIGMA-OCD and UKB separately). Values were selected to minimize the average Mean Squared Error (MSE) and average Relative Reconstruction Error (RRE) between the original mean time series ($f(t)$) and the reconstructed signal ($\sum_{k=1}^{K} u_k(t)$) across all subjects.

The errors were computed as:

$$\overline{\text{MSE}} = \frac{1}{N}\sum_{j=1}^{N}\left[\frac{1}{S}\sum_{i=1}^{S}\left(f_j(t_i) - \sum_{k=1}^{K} u_{k,j}(t_i)\right)^2\right] \text{ (Eq. S8)}$$

$$\overline{\text{RRE}} = \frac{1}{N}\sum_{j=1}^{N}\left[\frac{\left\|f_j(t) - \sum_{k=1}^{K} u_{k,j}(t)\right\|_2}{\|f_j(t)\|_2}\right] \text{ (Eq. S9)}$$

where $S$ is the number of time points in the signal and $N$ is the number of subjects. This tuning process resulted in $\alpha$ = 100 and $\tau$ = 3.5 for both ENIGMA-OCD and UKB datasets (Supplementary Table S7).

##### 3.5. Frequency band cutoff computation from VMD output

Once the VMD algorithm converged, yielding $K = 4$ IMFs ($u_k(t)$) and their final center frequencies ($\omega_k$) for the mean time series, the frequency band cutoffs for each IMF were determined (Supplementary Table S8). This process involved:

1. **Fourier transform:** The Fast Fourier Transform (FFT) was computed for each IMF, $u_k(t)$, to obtain its spectrum $U_k(\omega)$.
2. **Power spectrum:** The power spectrum for each IMF was calculated as $|U_k(\omega)|^2$.
3. **Thresholding:** A power threshold was applied to identify the significant frequency components. This threshold was set to 5% of the maximum power in the spectrum of that specific IMF.
4. **Frequency support:** The analysis was restricted to positive frequencies up to the Nyquist frequency ($f_s/2$, where $f_s$ is the sampling frequency equal to $1/\text{TR}$). Frequencies within this positive range whose power exceeded the threshold were considered part of the IMF's significant frequency support.
5. **Cutoff determination:** For each IMF, the lower bound ($f_{\text{min}}$) of its frequency band was defined as the minimum frequency in its significant support, and the higher bound ($f_{\text{max}}$) as the maximum frequency. These bounds were constrained to be non-negative and not to exceed the Nyquist frequency. This resulted in four pairs of cutoffs: ($f_{\text{imf1,lb}}$, $f_{\text{imf1,hb}}$), ..., ($f_{\text{imf4,lb}}$, $f_{\text{imf4,hb}}$).

##### 3.6. Extraction of frequency band time series

Using the subject-specific frequency cutoffs (e.g., [$f_{\text{imf1,lb}}$, $f_{\text{imf1,hb}}$], [$f_{\text{imf2,lb}}$, $f_{\text{imf2,hb}}$], etc.) determined from the mean time series, the original BOLD time series for each of the 304 ROIs were filtered to extract the activity within each of the four frequency bands. This was achieved by applying a bandpass filter for each of the four defined frequency ranges.

1. **Filter design:** A Butterworth bandpass filter of order 4 was designed for each of the four frequency bands ([$f_{lb}$, $f_{hb}$]) derived from the VMD output. The cutoff frequencies for the filter were normalized by the Nyquist frequency.
2. **Filtering:** The $filtfilt$ function from the scipy package[11] was used to apply the Butterworth filter to each ROI's time series. This zero-phase filtering approach ensures that the phase of the signal is not distorted. This process resulted in four distinct filtered time series for each ROI, corresponding to the four VMD-derived frequency bands. These four sets of 304 ROI time series were then used as input to the MBBN model.

### 4. Self-supervised pretraining on UK Biobank data

##### 4.1 Pretraining procedure

Our pretraining procedure followed the original MBBN approach[8], but with signals decomposed into four frequency bands using Variational Mode Decomposition (VMD), a deviation from the original Lorentzian/multifractal 3-band approach. Pretraining was conducted via Masked Signal Modeling (MSM), where the model must reconstruct missing data segments based on spatiotemporal context.

- Spatial Masking: Instead of random masking, we targeted nodes with high communicability, as this approach showed the best performance in the original MBBN model[8].

- Temporal Masking: Consecutive time windows were zeroed out to force the model to learn long-range temporal dependencies.

##### 4.2. The choice of a pretraining model setting

To identify the best performing pretrained model, we systematically varied three masking hyperparameters to produce 64 distinct model instances:

1. Number of Masked ROIs: 150, 200, 250, or 290 (out of 304 total).
2. Temporal Window Width: 10, 20, 30, or 40 time points.
3. Window Interval: 1, 2, 3, or 4 window widths apart.

The optimal pretrained model was identified via the two-tier selection process:

**Tier 1:** All 64 pretrained candidates were finetuned on a fixed training/validation split. In this stage, finetuning hyperparameters were held constant across all models. The top 5 models were selected based on the highest validation Area Under the Receiver Operating Characteristic curve (AUROC).

**Tier 2:** For the 5 finalists, we conducted a grid search across 20 predefined hyperparameter settings. This total of 100 configurations was finetuned on the same fixed split to identify the single best-performing "Pretrained Model + Fine-tuning HP" combination.

Final pretraining and finetuning hyperparameters are listed in the Supplementary Table S9.

### 5. Hyperparameter tuning procedure for baselines

##### 5.1. Overall strategy

All baseline models were trained and evaluated on three independent data splits, each defined by a different random seed. To optimize performance while managing computational cost, hyperparameter tuning for each model was conducted using only the first data split. For each model, the hyperparameter combination that yielded the highest Area Under the Receiver Operating Characteristic Curve (AUROC) on the validation set of the first split was selected. This final, optimized configuration was then used for the definitive evaluation across all three random seeds to produce the final reported performance metrics.

##### 5.2. Model-specific tuning procedures

1. **Support Vector Machine (SVM):** The SVM classifier was implemented using scikit-learn (v1.4.0)[12]. We first performed a search to identify the optimal kernel from a set of [linear, rbf, poly, sigmoid]. With the best-performing kernel selected, we then conducted a search over the regularization parameter C from a range of [1e-3, 1e-2, 0.1, 1, 10, 100, 1000].
2. **XGBoost:** The XGBoost model was implemented using the xgboost library (v2.0.3)[13]. We performed a comprehensive grid search over five key hyperparameters: max_depth [3, 6, 10], min_child_weight [1, 4, 7], gamma [0.0, 0.1, 0.4], learning_rate [0.05, 0.10, 0.30], and colsample_bytree [0.6, 0.8, 0.9].
3. **BNT and BolT:** These custom Transformer models, implemented in PyTorch (v2.7.1)[14], were tuned via a grid search. For BNT, we tuned learning_rate [1e-4, 3e-4, 5e-5], learning rate scheduler mode [cos, step], batch_size [8, 16, 32], and weight_decay [0, 1e-4, 1e-3]. For BolT, we tuned batch_size [8, 16, 32], learning_rate [1e-4, 2e-4], and weight_decay [0, 1e-4].
4. **Brain LM:** We evaluated several versions of the pretrained Brain LM model from Hugging Face (13M, 111M, 650M parameters), as well as a version trained from scratch. As our data contains 304 regions of interest (ROIs), a linear projection layer was added to map the input to the 434 ROIs expected by the pretrained models. Each model was fine-tuned over a range of learning rates specific to its size to identify the optimal configuration.
5. **Vanilla BERT:** This model, implemented using the transformers library (v4.54.0)[15], was tuned using the Optuna framework[16] for 20 trials. We optimized the initial learning rate, weight decay, and the gamma and step size parameters for the learning rate scheduler.

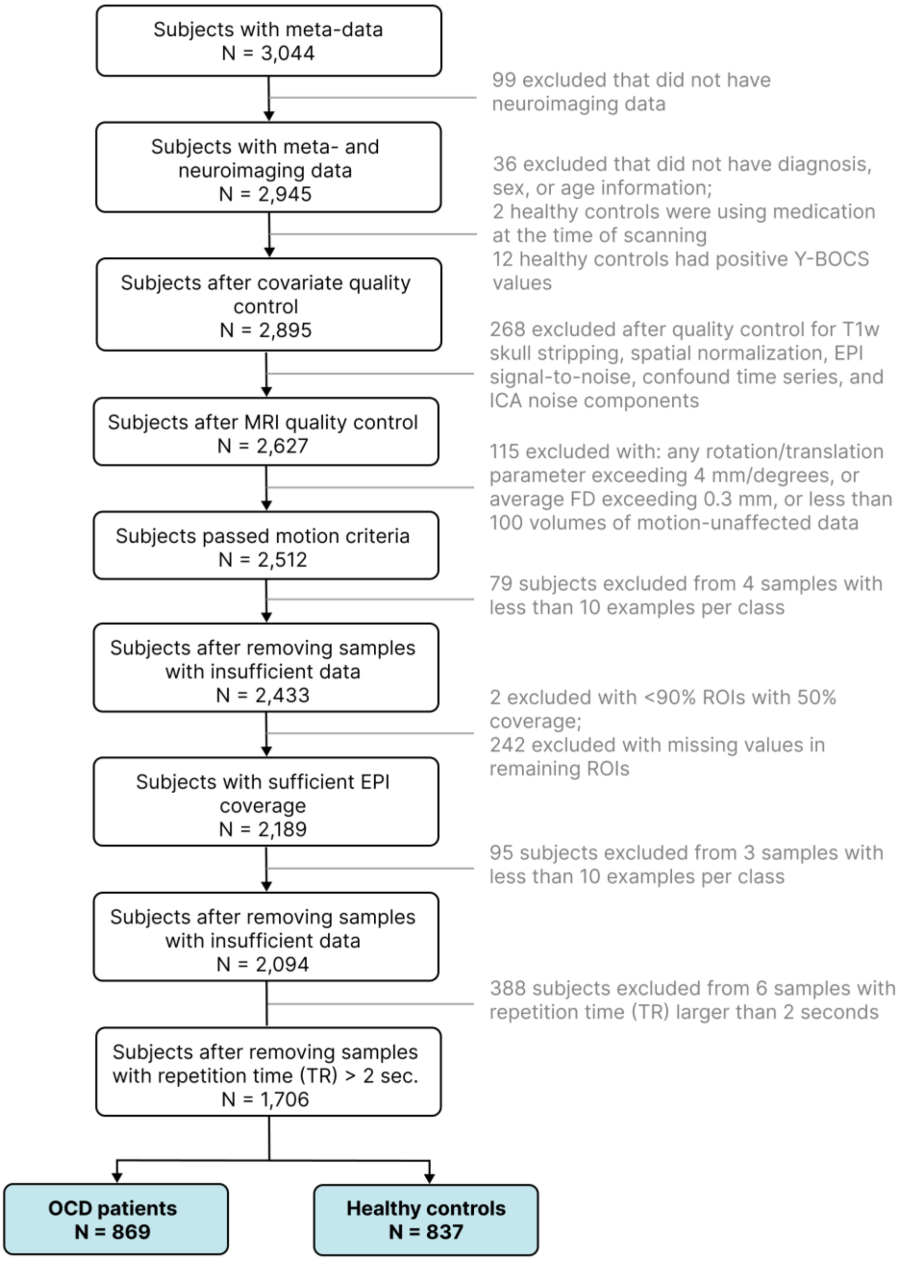


**Supplementary Figure S1. Study flowchart for participant selection and exclusion in the ENIGMA-OCD dataset. Abbreviations:** ENIGMA-OCD: Enhancing Neuro Imaging Genetics through Meta Analysis Obsessive-Compulsive Disorder, OCD: obsessive-compulsive disorder, Y-BOCS: Yale-Brown Obsessive-Compulsive Scale, MRI: magnetic resonance imaging, EPI: echo planar imaging, FD: framewise displacement, ICA: independent component analysis, T1w: T1-weighted, ROI: region of interest.

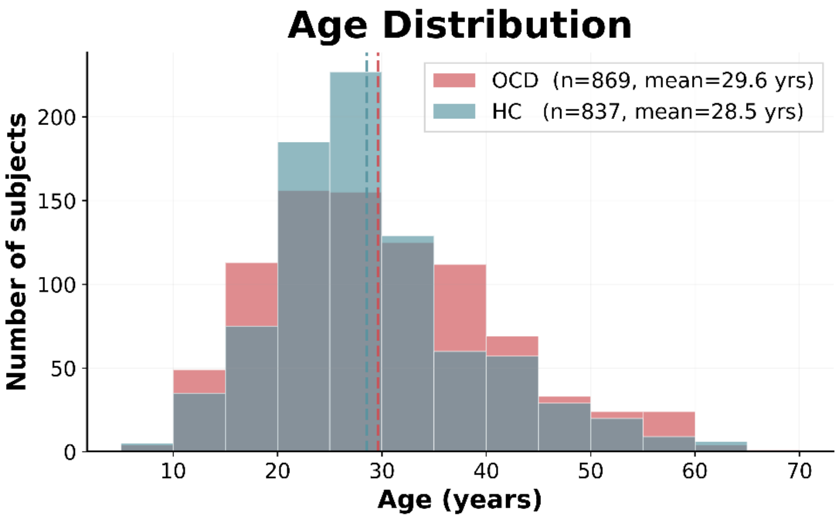


**Supplementary Figure S2. Age distributions in the OCD and healthy control (HC) groups.** Histograms show highly overlapping age distributions between groups, with similar mean ages (OCD: 29.6 years; HC: 28.5 years), supporting that age was well-balanced across the sample.

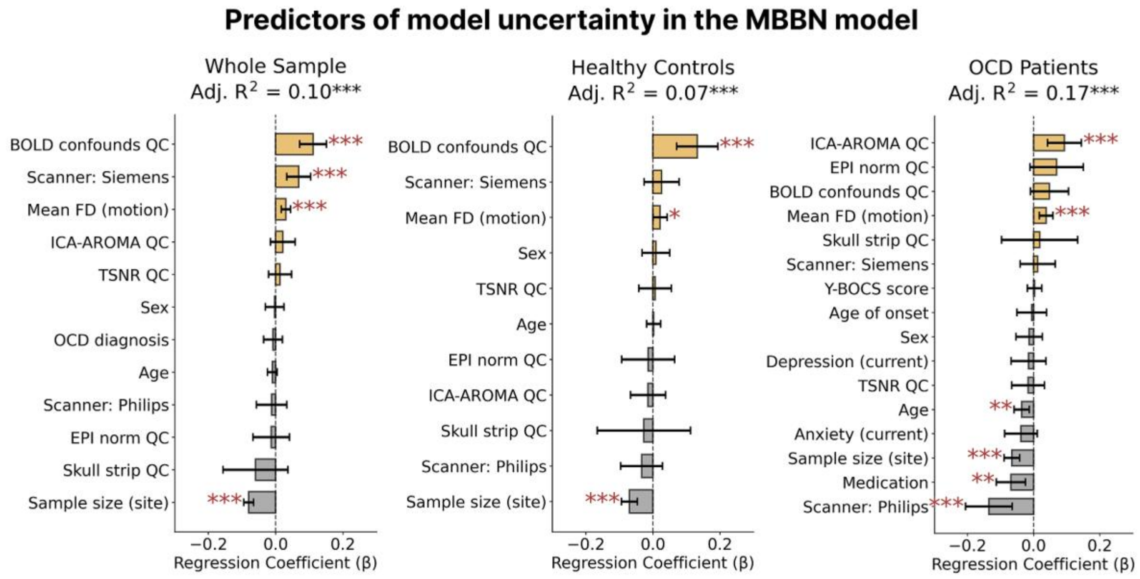


**Supplementary Figure S3. Uncertainty predictors in the from-scratch MBBN model, examined separately for the whole sample, healthy controls, and OCD patients.** Positive coefficients indicate factors associated with higher uncertainty. Quality control (QC) variables are binary-coded quality ratings (0 = good, 1 = uncertain). Significance levels: $p < .05$*, $p < .01$**, $p < .001$***. **Abbreviations:** MBBN: Multi-Band Brain Net, OCD: obsessive-compulsive disorder, BOLD: blood-oxygen-level-dependent, ICA-AROMA: independent component analysis-based automatic removal of motion artifacts, TSNR: temporal signal-to-noise ratio, FD: framewise displacement, EPI: echo planar imaging, Y-BOCS: Yale-Brown obsessive-compulsive scale, QC: quality control.

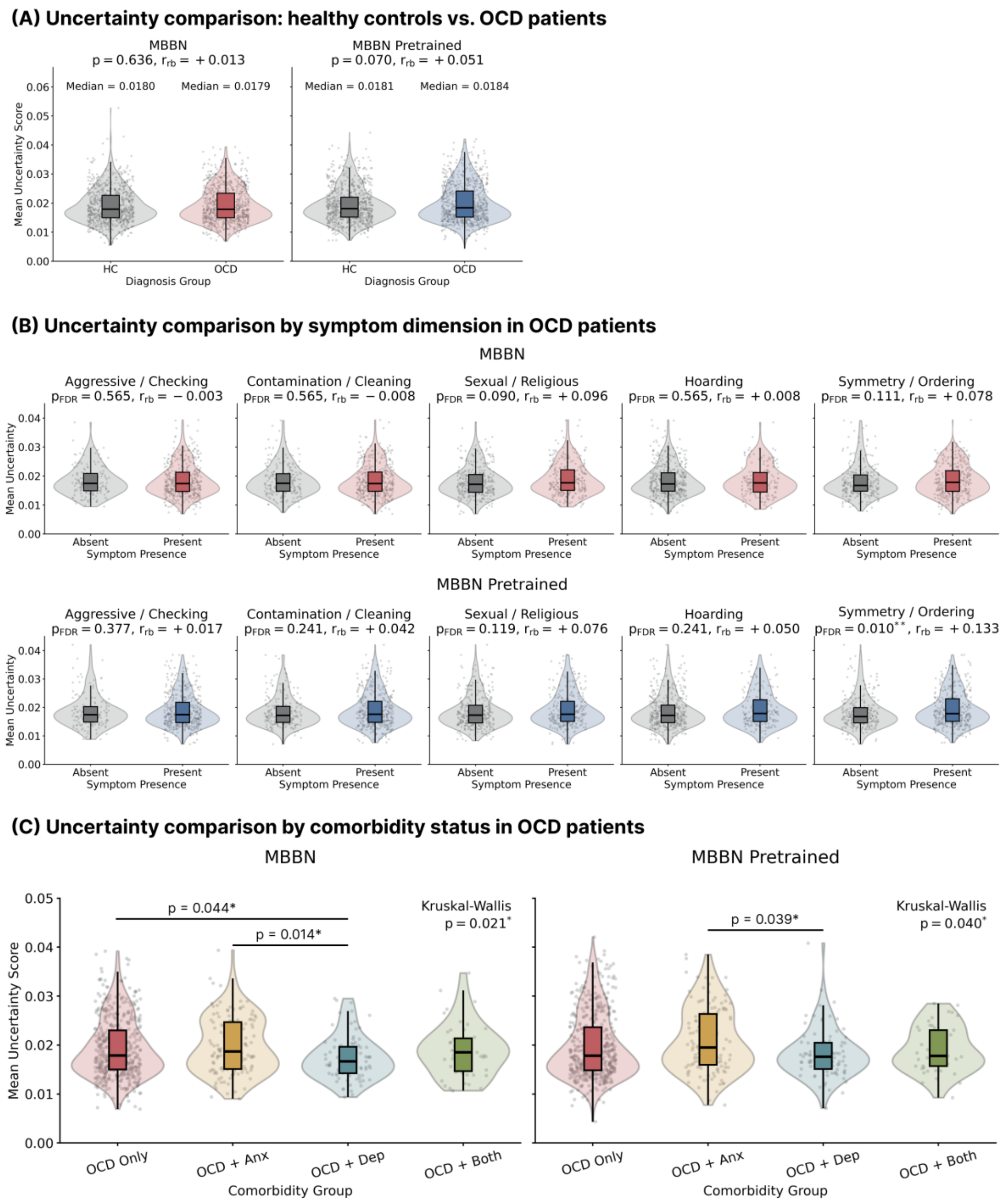


**Supplementary Figure S4. Association of model uncertainty with clinical phenotypes in OCD.** Statistical tests are non-parametric with effect sizes reported as rank-biserial correlation ($r_{rb}$). Asterisks indicate significance levels: $p < .05$*, $p < .01$**, $p < .001$***. **(A)** Uncertainty score comparison revealed no significant differences between healthy controls (HC) and OCD patients (MBBN from scratch: $p = .636$, MBBN pretrained: $p = .070$). **(B)** Uncertainty scores by symptom dimension in OCD patients. Only symmetry/ordering dimension showed a significant difference in the pretrained model ($p_{\text{FDR}} = .010$). **(C)** Uncertainty scores by comorbidity status in OCD patients. For both models, the OCD + Anxiety group showed significantly higher uncertainty than the OCD + Depression group after multiple comparisons correction (MBBN from scratch: $p = .014$, MBBN pretrained: $p = .039$). **Abbreviations:** MBBN: Multi-Band Brain Net, OCD: obsessive-compulsive, $r_{rb}$: rank-biserial correlation, FDR: false discovery rate.

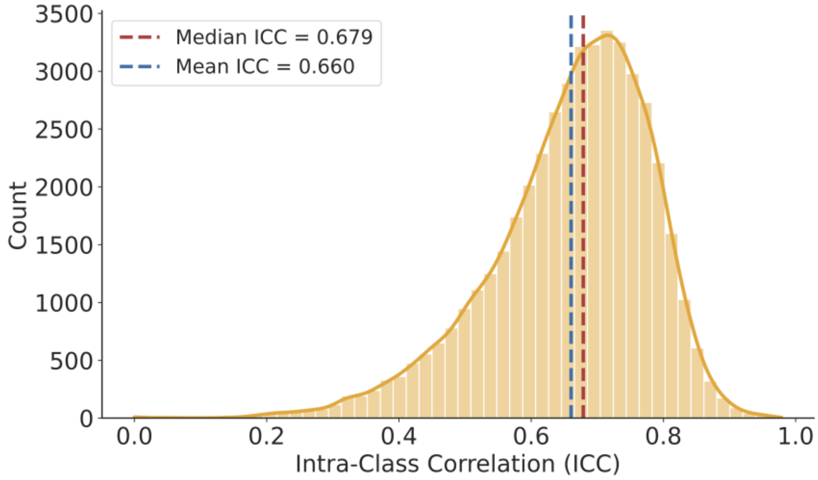


**Supplementary Figure S5. Quantification of site-level heterogeneity in functional connectivity using intra-class correlation (ICC) analysis.** Distribution of ICC values across 46,056 unique functional connectivity features (upper triangle of 304×304 connectivity matrix). ICC quantifies the proportion of variance attributable to site differences versus within-site (subject-level) variance, calculated using linear mixed-effects models with site as a random effect. Functional connectivity values were transformed using Fisher-z prior to analysis. A median ICC of 0.679 indicates high site-level heterogeneity and might explain the variability observed in leave-one-site-out cross-validation performance.

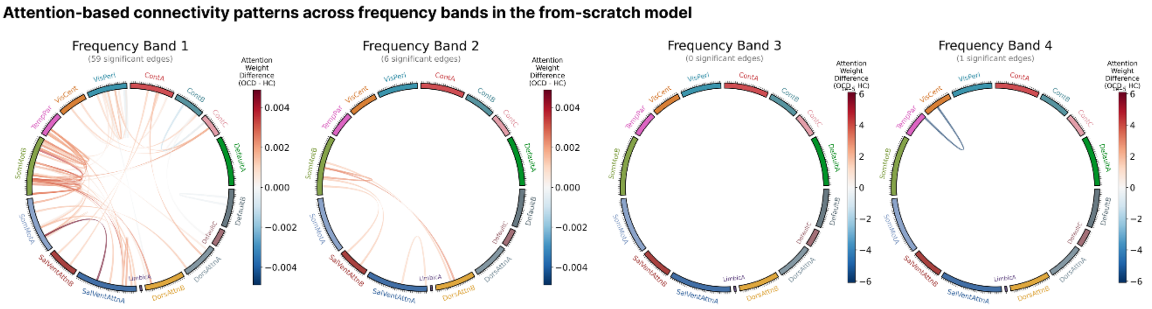


**Supplementary Figure S6. Attention-based functional connectivity (FC) patterns in the from-scratch model.** Significant FC edges ($p_{FDR} < .05$) between 304 ROIs from the Schaefer 400-parcel, 17-network atlas identified via attention weights. Blue edges indicate hypoconnectivity (OCD < HC); red edges indicate hyperconnectivity (OCD > HC). Band 1: 0.004-0.04 Hz, Band 2: 0.04-0.10 Hz, Band 3: 0.10-0.17 Hz, Band 4: 0.17-0.25 Hz. **Abbreviations:** OCD: obsessive-compulsive disorder, HC: healthy controls, ROI: region of interest, FDR: false discovery rate.

**Supplementary Table S1. Site-specific sample characteristics of the ENIGMA-OCD sample used in the analysis.** Demographic and clinical characteristics for each participating site, including sample size, age, sex distribution, education, and clinical measures (Y-BOCS scores, medication status). Values are presented as mean ± standard deviation for continuous variables and n (%) for categorical variables. **Abbreviations:** HC: healthy controls, OCD: obsessive-compulsive disorder, Y-BOCS: Yale-Brown Obsessive-Compulsive Scale, NA: not available, SD: standard deviation.

| | | | OCDs | | | | | | | HCs | | | |
|---|---|---|---|---|---|---|---|---|---|---|---|---|---|
| **Site** | **N** | **Age group** | **N** | **Age ± SD** | **Sex (male/ female)** | **Years of education ± SD** | **Medication use at time of scan (Yes/No/NA)** | **Age of onset (child/ adult/ NA)** | **Y-BOCS score ± SD** | **N** | **Age ± SD** | **Sex (male/ female)** | **Years of education ± SD** |
| **Amsterdam_VUmc** | 57 | adult | 29 | 37.21 ± 11.02 | 12/17 | 12.00 ± 3.02 | 27/0/2 | 18/9/2 | 20.52 ± 6.32 | 28 | 37.29 ± 9.50 | 13/15 | 12.68 ± 2.94 |
| **Bangalore_NIMHANS** | 358 | adult | 169 | 29.85 ± 6.74 | 87/82 | 13.00 ± 4.04 | 110/59/0 | 56/113/0 | 25.90 ± 6.21 | 189 | 27.07 ± 4.76 | 127/62 | 16.32 ± 2.87 |
| **Barcelona_HCPB** | 64 | pediatric + adult | 36 | 15.53 ± 2.13 | 19/17 | 9.50 ± 2.09 | 5/31/0 | 36/0/0 | 20.11 ± 8.03 | 28 | 15.61 ± 2.10 | 14/14 | 9.61 ± 2.10 |
| **Barcelone_Bellvitge/ ANTIGA_1.5T** | 137 | adult | 51 | 33.31 ± 9.23 | 25/26 | 12.76 ± 2.93 | 2/49/0 | 20/31/0 | 26.41 ± 4.87 | 86 | 33.12 ± 9.35 | 51/35 | NA |
| **Barcelone_Bellvitge/ PROV_1.5T** | 64 | adult | 43 | 37.16 ± 10.90 | 22/21 | 11.09 ± 3.19 | 3/40/0 | 16/27/0 | 21.88 ± 7.20 | 21 | 34.52 ± 7.85 | 11/10 | 13.30 ± 3.77 |
| **Barcelone_Bellvitge/ RESP_CBT_3T** | 57 | adult | 10 | 24.30 ± 3.92 | 9/1 | 12.70 ± 2.36 | 1/9/0 | 5/5/0 | 16.70 ± 8.53 | 47 | 28.55 ± 5.71 | 17/30 | 14.74 ± 2.98 |
| **Bergen** | 53 | adult | 29 | 30.03 ± 9.19 | 9/20 | 14.59 ± 2.23 | 20/9/0 | 11/16/2 | 26.31 ± 4.31 | 24 | 30.92 ± 10.51 | 7/17 | 14.38 ± 2.36 |
| **Braga_UMinho/ Braga_1.5T** | 46 | adult | 28 | 29.96 ± 10.45 | 10/18 | 11.82 ± 3.57 | 1/27/0 | 8/19/1 | 27.08 ± 4.90 | 18 | 32.22 ± 12.43 | 8/10 | 15.33 ± 3.68 |
| **Braga_UMinho/ Braga_1.5T_act** | 96 | adult | 44 | 27.93 ± 6.61 | 22/22 | 13.39 ± 2.92 | 0/44/0 | 22/21/1 | 25.88 ± 5.95 | 52 | 25.94 ± 5.11 | 17/35 | 15.71 ± 2.81 |
| **Braga_UMinho/ Braga_3T** | 59 | adult | 32 | 29.53 ± 11.02 | 14/18 | 13.22 ± 3.72 | 6/26/0 | 19/13/0 | 26.06 ± 4.66 | 27 | 30.41 ± 12.16 | 11/16 | 13.93 ± 4.02 |
| **Brazil** | 93 | adult | 59 | 38.34 ± 11.38 | 19/40 | 14.86 ± 4.75 | 23/34/2 | 44/10/5 | 29.21 ± 6.61 | 34 | 33.26 ± 11.31 | 15/19 | 16.42 ± 3.98 |
| **Cape_Town_UCT/ Skyra** | 39 | adult | 23 | 30.78 ± 9.44 | 10/13 | 14.48 ± 3.16 | 6/17/0 | 18/5/0 | 14.18 ± 13.09 | 16 | 30.81 ± 8.80 | 4/12 | 17.63 ± 2.87 |
| **Dresden** | 35 | pediatric + adult | 25 | 15.07 ± 1.95 | 11/14 | 8.56 ± 2.06 | 17/8/0 | 24/1/0 | 11.00 ± 8.68 | 10 | 15.40 ± 1.85 | 5/5 | 9.00 ± 1.83 |
| **Kyoto_KPU/ Kyoto3T** | 70 | adult | 34 | 33.05 ± 9.77 | 11/23 | 14.41 ± 2.27 | 34/0/0 | 9/25/0 | 22.50 ± 6.91 | 36 | 29.31 ± 7.59 | 18/18 | 15.19 ± 1.85 |
| **New_York** | 60 | adult | 49 | 29.49 ± 10.30 | 17/32 | 14.51 ± 1.82 | 14/35/0 | 0/0/49 | 23.84 ± 5.41 | 11 | 39.00 ± 10.80 | 5/6 | 13.55 ± 1.81 |
| **NYSPI_Columbia/ Adults** | 70 | adult | 38 | 29.29 ± 7.45 | 21/17 | 15.92 ± 2.65 | 37/0/1 | 22/15/1 | 13.60 ± 12.81 | 32 | 29.43 ± 7.75 | 15/17 | 16.69 ± 1.60 |
| **NYSPI_Columbia /Pediatric** | 34 | pediatric | 21 | 12.76 ± 3.06 | 10/11 | 8.00 ± 2.86 | 21/0/0 | 21/0/0 | 23.95 ± 5.23 | 13 | 13.08 ± 2.56 | 6/7 | 8.62 ± 3.64 |
| **UCLA** | 51 | adult | 32 | 31.28 ± 8.36 | 16/16 | 16.06 ± 2.20 | 22/9/1 | 30/1/1 | 24.66 ± 4.16 | 19 | 29.26 ± 10.23 | 10/9 | 15.68 ± 2.38 |
| **Vancouver_BCCHR** | 45 | pediatric + adult | 23 | 15.52 ± 2.32 | 9/14 | 9.07 ± 2.13 | 3/20/0 | 23/0/0 | 7.98 ± 8.83 | 22 | 13.92 ± 3.44 | 8/14 | 7.50 ± 3.60 |

| Site | N | Age group | OCDs | | | | | | | HCs | | | |
|---|---|---|---|---|---|---|---|---|---|---|---|---|---|
| | | | N | Age ± SD | Sex (male/ female) | Years of education ± SD | Medication use at time of scan (Yes/No/NA) | Age of onset (child/ adult/ NA) | Y-BOCS score ± SD | N | Age ± SD | Sex (male/ female) | Years of education ± SD |
| **Yale_Gruner** | 29 | pediatric | 13 | 14.35 ± 1.78 | 5/8 | NA | 6/7/0 | 13/0/0 | 26.31 ± 3.86 | 16 | 14.33 ± 1.92 | 7/9 | NA |
| **Yale_Pittinger/ HCP_Prisma** | 61 | adult | 35 | 30.91 ± 11.34 | 16/19 | 15.39 ± 2.05 | 23/11/1 | 0/0/35 | 13.00 ± 11.42 | 26 | 28.73 ± 8.02 | 15/11 | 16.65 ± 2.23 |
| **Yale_Pittinger/ HCP_Trio** | 43 | adult | 21 | 37.62 ± 11.86 | 5/16 | 14.67 ± 2.59 | 21/0/0 | 1/1/19 | 12.05 ± 13.61 | 22 | 33.64 ± 14.13 | 8/14 | 15.91 ± 2.27 |
| **Yale_Pittinger/ Yale_2014** | 85 | adult | 25 | 37.16 ± 13.79 | 14/11 | 15.36 ± 2.29 | 13/12/0 | 8/17/0 | 27.36 ± 5.92 | 60 | 32.27 ± 9.96 | 34/26 | 15.92 ± 3.14 |

**Supplementary Table S2. Comorbid disorders and symptom dimensions of the OCD group (N = 869) in the final ENIGMA-OCD sample.** Comorbid disorders include current and lifetime depression and anxiety. OCD symptom dimensions are based on the Yale-Brown Obsessive-Compulsive Scale (Y-BOCS) symptom checklist. Values represent counts for each category (no, yes, NA). **Abbreviations:** NA: not available, OCD: obsessive-compulsive disorder.

| Category | Item | No | Yes | NA |
|---|---|---|---|---|
| **Comorbid disorder** | Depression current | 652 | 135 | 82 |
| | Depression lifetime | 481 | 187 | 201 |
| | Anxiety current | 600 | 179 | 90 |
| | Anxiety lifetime | 483 | 185 | 201 |
| **OCD symptom dimension** | Aggressive / checking | 156 | 491 | 222 |
| | Contamination / cleaning | 217 | 430 | 222 |
| | Symmetry / ordering | 272 | 375 | 222 |
| | Sexual / religious | 353 | 294 | 222 |
| | Hoarding | 457 | 190 | 222 |

**Supplementary Table S3. Image acquisition parameters for the ENIGMA-OCD sample used in the analysis.** Site-specific acquisition parameters for structural and functional MRI scans, including scanner type, field strength, voxel size, repetition time (TR), echo time (TE), flip angle, and number of volumes. Study inclusion dates and publication DOIs are provided where available. **Abbreviations:** TR: repetition time, TE: echo time, NA: not available.

| **Site** | **N** | **Field strength (Tesla) and scanner type** | **STRUCTURAL SCAN PARAMS** | | | | **FUNCTIONAL SCAN PARAMS** | | | | | **Study inclusion start date (YYYY-MM-DD)** | **Study inclusion end date (YYYY-MM-DD)** | **DOI to published paper (if available)** |
|---|---|---|---|---|---|---|---|---|---|---|---|---|---|---|
| | | | **Voxel-size (X, Y, Z) (mm)** | **TR (s)** | **TE (ms)** | **Flip angle (°)** | **Voxel-size (X ,Y, Z) (mm)** | **TR (s)** | **TE (ms)** | **Flip angle (°)** | **N Volumes** | | | |
| **Amsterdam_VUmc** | 57 | 3T GE Signa HDxt | 1, 0.98, 0.98 | NA | NA | NA | 3.3, 3.3, 3 | 1.8 | 35 | 80 | 200 | 2008-01-01 | 2014-01-01 | 10.1080/15622975.2017.1353132 |
| **Bangalore_NIMHANS** | 358 | 3T Siemens Skyra | 1, 1, 1 | 1.9 | 2.43 | 9 | 3, 3, 3 | 2 | 30 | 78 | 153-303 | 2010 | NA | NA |
| **Barcelona_HCPB** | 64 | 3T TrioTim | 1, 0.94, 0.94 | 2.3 | 3.01 | 9 | 3, 3, 4 | 2 | 29 | 80 | 240 | 2007-07-01 | 2013-06-30 | NA |
| **Barcelone_Bellvitge/ANTIGA_1.5T** | 137 | 1.5T GE Signa Excite | 1.17, 1.17, 1.2 | 11.8 | 4.2 | 15 | 3.75, 3.75, 5 | 2 | 50 | 90 | 120 | 2006-07-19 / 2009-02-04 | 2007-12-21 / 2010-03-31 | 10.1016/j.biopsych.2012.10.006 |
| **Barcelone_Bellvitge/PROV_1.5T** | 64 | 1.5T GE Signa Excite | 1.17, 1.17, 1.2 | 11.8 | 4.2 | 15 | 3.75, 3.75, 5 | 2 | 50 | 90 | 120 | 2011-05-12 | 2013-02-19 | 10.1017/S0033291717002288 |
| **Barcelone_Bellvitge/RESP_CBT_3T** | 57 | 3T Philips Ingenia | 0.75, 0.75, 0.75 | 10.68 | 4.96 | 8 | 3, 3, 3 | 2 | 25 | 90 | 240 | 2016-12-14 | continued | unpublished |
| **Bergen** | 53 | 3T GE Discovery MR750 | 1, 1, 1 | 7 | 3 | 12 | 3.44, 3.44, 3.3 | 1.8 | 30 | 80 | 160 | 2015-08-24 | 2017-12-11 | 10.1016/j.bpsc.2020.01.007 |
| **Braga_UMinho/ Braga_1.5T** | 46 | 1.5T Siemens Magnetom Avanto | 1, 1, 1 | 2.73 | 3.48 | 7 | 3.5, 3.5, 3.5 | 2 | 30 | 90 | 180 | 2012-05-23 | 2017-04-07 | 10.1016/j.pscychresns.2019.06.008 |
| **Braga_UMinho/ Braga_1.5T_act** | 96 | 1.5T Siemens Magnetom Avanto | 1, 1, 1 | 2.73 | 3.48 | 7 | 3.5, 3.5, 3.5 | 2 | 30 | 90 | 180 | 2012-05-23 | 2017-04-07 | 10.1016/j.pscychresns.2019.06.008 |
| **Braga_UMinho/ Braga_3T** | 59 | 3T Siemens Verio | 1, 1, 1 | 2.42 | 4.12 | 9 | 2, 2, 2 | 1 | 27 | 62 | 720 | 2019-05-08 | 2020-11-04 | NA |
| **Brazil** | 93 | 3T Philips Medical Systems MRI Scanner | 1, 1, 1 | 7 | 3.2 | 8 | 3, 3, 3 | 2 | 30 | 80 | 130 | 2014-08-19 | 2017-11-27 | NA |
| **Cape_Town_UCT/ Skyra** | 39 | 3T Siemens Skyra | 1, 1, 1 | 2.53 | 1.69 | 7 | 3.75, 3.75, 5 | 1.73 | 27 | 70 | 298 | 28-05-2015 | NA | NA |
| **Dresden** | 35 | 3T Siemens MAGNETOM Prisma | 0.8, 0.8, 0.8 | NA | NA | NA | 2, 2, 2 | 0.8 | 37 | 52 | 144 | NA | NA | NA |
| **Kyoto_KPU/ Kyoto3T** | 70 | 3T Philips Achieva TX | 1, 1, 1 | 7.1 | 3.3 | 10 | 3, 3, 3 | 2 | 30 | 80 | 200 | 2010-08-14 | 2012-03-09 | 10.1016/j.euroneuro.2015.08.017 |
| **New_York** | 60 | 3T Siemens NKI TRIOTIM | 0.8, 0.8, 0.8 | 2.4 | 2.01 | 8 | 2.1, 2.1, 2.1 | 1 | 25.4 | 60 | 480 | 2018-04-30 | 2020-03-10 | NA |

| Site | N | Field strength (Tesla) and scanner type | STRUCTURAL SCAN PARAMS | | | | FUNCTIONAL SCAN PARAMS | | | | | Study inclusion start date (YYYY-MM-DD) | Study inclusion end date (YYYY-MM-DD) | DOI to published paper (if available) |
|---|---|---|---|---|---|---|---|---|---|---|---|---|---|---|
| | | | Voxel-size (X, Y, Z) (mm) | TR (s) | TE (ms) | Flip angle (°) | Voxel-size (X ,Y, Z) (mm) | TR (s) | TE (ms) | Flip angle (°) | N Volumes | | | |
| **NYSPI_Columbia/ Adults** | 70 | 3T GE MR750 | 0.8, 0.8, 0.8 | 7.86 | 3.11 | 12 | 2, 2, 2 | 0.85 | 25 | 60 | 544 | 2015-03-03 | 2019-11-18 | doi: 10.1038/s41386-020-00929-9 |
| **NYSPI_Columbia/ Pediatric** | 34 | 3T GE MR750 | 0.8, 0.8, 0.8 | 7.86 | 3.11 | 12 | 2, 2, 2 | 0.85 | 25 | 60 | 544 | 2015-04-25 | 2018-05-21 | doi: 10.1038/s41386-020-0613-3 doi: 10.1002/da.23187 |
| **UCLA** | 51 | 3T Siemens Trio | 1, 1, 1 | 1.9 | 3.26 | 9 | 3, 3, 3 | 2 | 25 | 78 | 208 | 2011-07-01 | 2015-06-30 | doi: 10.1073/pnas.1716686115; doi: 10.3389/fpsyt.2015.00074; doi: 10.1038/tp.2017.192; DOI: 10.2147/PRBM.S75106; doi: 10.1007/s11682-020-00358-8; doi: 10.1038/npp.2017.249 |
| **Vancouver_BCCHR** | 45 | 3T GE Discovery MR750 | 1, 1, 1 | 8.15 | 3.17 | 10 | 3, 3, 4 | 2 | 25 | 90 | 150 | 2017 | 2019 | NA |
| **Yale_Gruner** | 29 | 3T GE Signa | 0.98, 0.98, 1 | NA | NA | NA | 3.75, 3.75, 3 | 2 | 30 | 80 | 150 | NA | NA | 10.1002/hbm.22551 |
| **Yale_Pittinger/ HCP_Prisma** | 61 | 3T Prisma | 0.8, 0.8, 0.8 | NA | NA | NA | 2, 2, 2 | 0.8 | 37 | 52 | 500 | NA | NA | NA |
| **Yale_Pittinger/ HCP_Trio** | 43 | 3T Siemens Trio | 0.8, 0.8, 0.8 | NA | NA | NA | 2.5, 2.5, 2.5 | 0.7 | 31 | 55 | 500 | NA | NA | NA |
| **Yale_Pittinger/ Yale_2014** | 85 | 3T Siemens Trio | 1, 1, 1 | 2.53 | 3.66 | 7 | 3.44, 3.44, 4 | 2 | 25 | 85 | 300 | NA | NA | 10.1016/j.biopsych.2013.10.021 |

**Supplementary Table S4. OCD classification performance of MBBN and baseline models under different frequency filtering conditions.** Performance metrics (AUROC, Balanced Accuracy, Sensitivity, Specificity) for OCD vs. healthy control classification. Values represent mean ± standard deviation across random seeds. Models are categorized by input data type: functional connectivity (FC) or time series (TS). The highest performance for each metric is shown in **bold**, and the second highest is <u>underscored</u>. **Abbreviations:** AUROC: area under the receiver operating characteristic curve, Bal. Acc.: balanced accuracy, FC: functional connectivity, TS: time series, MBBN: Multi-Band Brain Network, OCD: obsessive-compulsive disorder, SD: standard deviation.

| Model | AUROC ± SD | Bal. Acc. ± SD | Sensitivity ± SD | Specificity ± SD |
|---|---|---|---|---|
| ***Low-pass filtered data (~0.25 Hz, main results)*** | | | | |
| **SVM (FC)** | 64.21 ± 2.06 | 59.92 ± 2.34 | 62.23 ± 1.96 | 57.62 ± 4.64 |
| **XGBoost (FC)** | 59.33 ± 3.14 | 56.23 ± 3.11 | 57.85 ± 3.99 | 54.60 ± 4.90 |
| **BNT (FC)** | <u>65.03 ± 2.93</u> | **60.60 ± 2.75** | 63.14 ± 5.91 | 58.05 ± 6.89 |
| **BolT (TS)** | 63.23 ± 2.81 | 58.93 ± 2.33 | 60.66 ± 8.06 | 57.21 ± 8.65 |
| **Brain LM (TS)** | 57.12 ± 2.77 | 53.81 ± 2.54 | <u>65.15 ± 10.55</u> | 42.46 ± 10.75 |
| **Vanilla BERT (TS)** | 60.83 ± 2.93 | 56.47 ± 3.39 | 35.82 ± 21.52 | **77.13 ± 15.85** |
| **MBBN from scratch (TS)** | **65.26 ± 3.85** | <u>60.36 ± 4.12</u> | **66.86 ± 16.87** | 53.87 ± 12.47 |
| **MBBN pretrained (TS)** | 63.93 ± 3.90 | 58.52 ± 3.87 | 47.79 ± 23.69 | <u>69.24 ± 22.63</u> |
| ***Band-pass filtered data (0.01-0.1 Hz, control analysis)*** | | | | |
| **MBBN from scratch (TS)** | 62.46 ± 3.88 | 57.75 ± 2.82 | 40.05 ± 24.33 | 75.44 ± 19.80 |
| **MBBN pretrained (TS)** | 62.20 ± 4.47 | 58.82 ± 5.67 | 53.07 ± 7.03 | 64.56 ± 10.21 |

**Supplementary Table S5. OCD classification performance on unfiltered fMRI data.** Performance metrics (AUROC, Balanced Accuracy, Sensitivity, Specificity) for OCD vs. healthy control classification. Values represent mean ± standard deviation across three random seeds. Models are categorized by input data type: functional connectivity (FC) or time series (TS). The highest performance for each metric is shown in **bold**, and the second highest is <u>underscored</u>. **Abbreviations:** AUROC: area under the receiver operating characteristic curve, Bal. Acc.: balanced accuracy, FC: functional connectivity, TS: time series, MBBN: Multi-Band Brain Network, OCD: obsessive-compulsive disorder, SD: standard deviation.

| Model | AUROC ± SD | Bal. Acc. ± SD | Sensitivity ± SD | Specificity ± SD |
|---|---|---|---|---|
| **SVM (FC)** | 62.47 ± 2.16 | <u>58.75 ± 0.65</u> | **61.11 ± 2.25** | 56.39 ± 0.96 |
| **XGBoost (FC)** | 59.24 ± 1.81 | 56.96 ± 1.19 | 60.26 ± 3.18 | 53.67 ± 1.29 |
| **BNT (FC)** | **65.25 ± 1.78** | **61.36 ± 1.92** | <u>60.82 ± 4.53</u> | 61.90 ± 7.37 |
| **BolT (TS)** | 61.87 ± 3.34 | 57.31 ± 1.48 | 54.27 ± 5.98 | 60.33 ± 8.24 |
| **Brain LM (TS)** | 56.70 ± 2.99 | 54.20 ± 0.82 | 56.62 ± 17.78 | 51.78 ± 16.14 |
| **Vanilla BERT (TS)** | 61.72 ± 3.97 | 58.18 ± 3.12 | 52.27 ± 14.14 | 64.08 ± 12.79 |
| **MBBN from scratch (TS)** | <u>63.41 ± 4.96</u> | 58.20 ± 4.17 | 36.67 ± 16.10 | **79.72 ± 11.39** |
| **MBBN pretrained (TS)** | 62.32 ± 2.98 | 57.22 ± 4.05 | 36.87 ± 7.24 | <u>77.56 ± 14.33</u> |

**Supplementary Table S6. Classification performance across different numbers of frequency bands: A control analysis justifying the selection of four frequency bands.** Performance metrics for OCD vs healthy control classification using the MBBN from-scratch model trained with different numbers of frequency bands (1-5). Values represent mean ± standard deviation across three random seeds. Four frequency bands shown in **bold** yielded the highest AUROC (67.36 ± 2.89) and balanced accuracy (61.69 ± 4.49), confirming the optimal number of bands for decomposition. The reported AUROC and SD differ from the main text, because control analyses were performed on three data splits instead of 10, because of the high computational cost. **Abbreviations:** AUROC: area under the receiver operating characteristic curve, MBBN: Multi-Band Brain Network, OCD: obsessive-compulsive disorder, SD: standard deviation.

| Number of frequency bands | AUROC ± SD | Bal. Acc. ± SD |
|---|---|---|
| 1 | 61.12 ± 2.81 | 56.80 ± 1.85 |
| 2 | 60.26 ± 7.28 | 56.39 ± 3.07 |
| 3 | 59.79 ± 6.88 | 56.07 ± 2.74 |
| **4** | **67.36 ± 2.89** | **61.69 ± 4.49** |
| 5 | 65.57 ± 5.10 | 61.37 ± 2.12 |

**Supplementary Table S7. Optimized Variational Mode Decomposition (VMD) parameters for the ENIGMA-OCD and UK Biobank datasets.** VMD parameters were optimized to minimize reconstruction error, ensuring the sum of extracted Intrinsic Mode Functions (IMFs) closely matched the original rs-fMRI signal. Parameters include the bandwidth constraint (alpha) and the Lagrangian multiplier update rate (tau). **Abbreviations:** VMD: variational mode decomposition, IMF: intrinsic mode function, rs-fMRI: resting-state functional magnetic resonance imaging.

| **Parameters** | | **ENIGMA-OCD** | | **UKB** | |
|---|---|---|---|---|---|
| **Alpha (α)** | **Tau (τ)** | **MSE** | **RRE** | **MSE** | **RRE** |
| 100 | 0.5 | 1.01e-03 | 2.36e-02 | 4.98e-10 | 1.53e-05 |
| 100 | 1 | 1.01e-03 | 2.36e-02 | 1.52e-10 | 9.19e-06 |
| 100 | 1.5 | 1.01e-03 | 2.36e-02 | 8.73e-11 | 7.27e-06 |
| 100 | 2 | 1.01e-03 | 2.36e-02 | 6.43e-11 | 6.37e-06 |
| 100 | 2.5 | 1.01e-03 | 2.36e-02 | 5.37e-11 | 5.85e-06 |
| 100 | 3 | 1.01e-03 | 2.36e-02 | 4.79e-11 | 5.53e-06 |
| **100** | **3.5** | **1.01e-03** | **2.36e-02** | **4.44e-11** | **5.31e-06** |
| 200 | 0.5 | 1.33e-03 | 2.78e-02 | 1.15e-09 | 1.94e-05 |
| 200 | 1 | 1.06e-03 | 2.43e-02 | 3.20e-10 | 1.13e-05 |
| 200 | 1.5 | 1.02e-03 | 2.37e-02 | 1.64e-10 | 8.70e-06 |
| 200 | 2 | 1.01e-03 | 2.36e-02 | 1.08e-10 | 7.43e-06 |
| 200 | 2.5 | 1.01e-03 | 2.36e-02 | 8.16e-11 | 6.69e-06 |
| 200 | 3 | 1.01e-03 | 2.36e-02 | 6.73e-11 | 6.21e-06 |
| 200 | 3.5 | 1.01e-03 | 2.36e-02 | 5.87e-11 | 5.88e-06 |
| 300 | 0.5 | 4.83e-03 | 5.74e-02 | 1.57e-09 | 1.83e-05 |
| 300 | 1 | 2.26e-03 | 3.61e-02 | 4.46e-10 | 1.11e-05 |
| 300 | 1.5 | 1.50e-03 | 2.90e-02 | 2.26e-10 | 8.75e-06 |
| 300 | 2 | 1.17e-03 | 2.56e-02 | 1.44e-10 | 7.54e-06 |
| 300 | 2.5 | 1.07e-03 | 2.43e-02 | 1.07e-10 | 6.84e-06 |
| 300 | 3 | 1.02e-03 | 2.38e-02 | 8.52e-11 | 6.37e-06 |
| 300 | 3.5 | 1.01e-03 | 2.36e-02 | 7.22e-11 | 6.04e-06 |
| 400 | 0.5 | 1.41e-02 | 1.05e-01 | 2.39e-09 | 2.04e-05 |
| 400 | 1 | 7.03e-03 | 6.72e-02 | 5.91e-10 | 1.07e-05 |
| 400 | 1.5 | 3.99e-03 | 4.84e-02 | 2.97e-10 | 8.50e-06 |
| 400 | 2 | 2.68e-03 | 3.88e-02 | 1.85e-10 | 7.40e-06 |
| 400 | 2.5 | 1.81e-03 | 3.18e-02 | 1.31e-10 | 6.74e-06 |
| 400 | 3 | 1.33e-03 | 2.75e-02 | 1.01e-10 | 6.31e-06 |
| 400 | 3.5 | 1.15e-03 | 2.54e-02 | 8.46e-11 | 6.00e-06 |
| 500 | 0.5 | 2.64e-02 | 1.49e-01 | 4.52e-09 | 3.19e-05 |
| 500 | 1 | 1.57e-02 | 1.09e-01 | 6.87e-10 | 1.10e-05 |
| 500 | 1.5 | 9.93e-03 | 8.17e-02 | 3.29e-10 | 8.37e-06 |
| 500 | 2 | 6.99e-03 | 6.52e-02 | 2.12e-10 | 7.34e-06 |
| 500 | 2.5 | 4.73e-03 | 5.19e-02 | 1.52e-10 | 6.71e-06 |
| 500 | 3 | 2.96e-03 | 4.11e-02 | 1.19e-10 | 6.29e-06 |
| 500 | 3.5 | 2.24e-03 | 3.51e-02 | 9.76e-11 | 5.98e-06 |
| 600 | 0.5 | 4.14e-02 | 1.89e-01 | 1.14e-08 | 5.75e-05 |
| 600 | 1 | 2.78e-02 | 1.51e-01 | 1.07e-09 | 1.37e-05 |
| 600 | 1.5 | 1.94e-02 | 1.21e-01 | 4.31e-10 | 8.81e-06 |
| 600 | 2 | 1.37e-02 | 9.78e-02 | 2.52e-10 | 7.44e-06 |
| 600 | 2.5 | 1.00e-02 | 8.00e-02 | 1.96e-10 | 6.78e-06 |
| 600 | 3 | 7.54e-03 | 6.69e-02 | 1.46e-10 | 6.34e-06 |
| 600 | 3.5 | 5.15e-03 | 5.45e-02 | 1.15e-10 | 6.02e-06 |
| 700 | 0.5 | 5.96e-02 | 2.29e-01 | 3.04e-08 | 9.95e-05 |
| 700 | 1 | 4.15e-02 | 1.87e-01 | 2.00e-09 | 2.00e-05 |
| 700 | 1.5 | 3.11e-02 | 1.59e-01 | 5.39e-10 | 1.00e-05 |
| 700 | 2 | 2.35e-02 | 1.33e-01 | 2.83e-10 | 7.69e-06 |
| 700 | 2.5 | 1.77e-02 | 1.12e-01 | 1.90e-10 | 6.81e-06 |
| 700 | 3 | 1.37e-02 | 9.56e-02 | 1.42e-10 | 6.33e-06 |
| 700 | 3.5 | 9.99e-03 | 7.94e-02 | 1.16e-10 | 6.02e-06 |
| 800 | 0.5 | 7.65e-02 | 2.60e-01 | 6.82e-08 | 1.57e-04 |
| 800 | 1 | 5.75e-02 | 2.23e-01 | 4.31e-09 | 3.16e-05 |
| 800 | 1.5 | 4.52e-02 | 1.95e-01 | 9.13e-10 | 1.29e-05 |
| 800 | 2 | 3.54e-02 | 1.69e-01 | 3.92e-10 | 8.52e-06 |
| 800 | 2.5 | 2.86e-02 | 1.49e-01 | 2.57e-10 | 7.10e-06 |
| 800 | 3 | 2.30e-02 | 1.29e-01 | 1.89e-10 | 6.48e-06 |
| 800 | 3.5 | 1.73e-02 | 1.10e-01 | 1.46e-10 | 6.10e-06 |
| 900 | 0.5 | 9.46e-02 | 2.90e-01 | 2.39e-07 | 2.32e-04 |
| 900 | 1 | 7.32e-02 | 2.53e-01 | 9.41e-09 | 4.94e-05 |
| 900 | 1.5 | 6.02e-02 | 2.28e-01 | 1.71e-09 | 1.84e-05 |
| 900 | 2 | 5.02e-02 | 2.05e-01 | 5.55e-10 | 1.03e-05 |
| 900 | 2.5 | 3.94e-02 | 1.79e-01 | 2.95e-10 | 7.74e-06 |
| 900 | 3 | 3.16e-02 | 1.58e-01 | 1.98e-10 | 6.69e-06 |
| 900 | 3.5 | 2.78e-02 | 1.44e-01 | 1.50e-10 | 6.17e-06 |
| 1,000 | 0.5 | 1.11e-01 | 3.18e-01 | 3.62e-07 | 3.19e-04 |
| 1,000 | 1 | 9.28e-02 | 2.87e-01 | 1.88e-08 | 7.39e-05 |
| 1,000 | 1.5 | 7.78e-02 | 2.59e-01 | 3.48e-09 | 2.73e-05 |
| 1,000 | 2 | 6.65e-02 | 2.38e-01 | 1.03e-09 | 1.39e-05 |
| 1,000 | 2.5 | 5.27e-02 | 2.11e-01 | 4.71e-10 | 9.25e-06 |
| 1,000 | 3 | 4.56e-02 | 1.93e-01 | 2.82e-10 | 7.38e-06 |
| 1,000 | 3.5 | 3.84e-02 | 1.74e-01 | 2.01e-10 | 6.53e-06 |

**Supplementary Table S8. Frequency band cutoffs from variational mode decomposition (VMD) across all subjects in the ENIGMA-OCD sample.** Frequency band boundaries and bandwidths were computed for each subject using VMD with four intrinsic mode functions (IMFs). Values represent mean ± standard deviation across all subjects (N = 1,706). Lower and upper bounds indicate the frequency range for each band, and bandwidth represents the difference between upper and lower bounds. **Abbreviation:** SD: standard deviation.

| Frequency Band | Lower Bound ± SD (Hz) | Upper Bound ± SD (Hz) | Bandwidth ± SD (Hz) |
|---|---|---|---|
| Band 1 (lowest) | 0.0040 ± 0.0021 | 0.0409 ± 0.0114 | 0.0368 ± 0.0112 |
| Band 2 | 0.0424 ± 0.0105 | 0.1013 ± 0.0167 | 0.0588 ± 0.0120 |
| Band 3 | 0.1016 ± 0.0164 | 0.1726 ± 0.0166 | 0.0710 ± 0.0119 |
| Band 4 (highest) | 0.1720 ± 0.0157 | 0.2485 ± 0.0088 | 0.0766 ± 0.0187 |

**Supplementary Table S9. Training hyperparameters for MBBN models.** Hyperparameter values for pretraining (UK Biobank), from-scratch training (ENIGMA-OCD), and finetuning (ENIGMA-OCD) configurations. For pretraining, the selected configuration (250 masked hub ROIs, window size 20, interval rate 4) was chosen from 64 candidate configurations through a two-tier selection process. **Abbreviations:** MBBN: Multi-Band Brain Network, ROI: region of interest, AdamW: Adam with decoupled weight decay.

| Hyperparameter | Pretraining (UK Biobank) | From-Scratch (ENIGMA-OCD) | Finetuning (ENIGMA-OCD) |
|---|---|---|---|
| **Model Architecture** | | | |
| Number of ROIs | 304 | 304 | 304 |
| Number of attention heads | 8 | 8 | 8 |
| Transformer hidden layers | 8 | 8 | 8 |
| Transformer dropout rate | 0.3 | 0.3 | 0.3 |
| Sequence length (time points) | 464 | 464 | 464 |
| **Training Configuration** | | | |
| Batch size | 32 | 16 | 32 |
| Optimizer | AdamW | AdamW | AdamW |
| **Learning Rate Schedule** | | | |
| Learning rate policy | step | step | step |
| Initial learning rate | $3\times10^{-4}$ | $5\times10^{-5}$ | $5\times10^{-5}$ |
| Learning rate decay | 0.95 | 0.95 | 0.97 |
| Learning rate step | 4500 | 4000 | 3000 |
| Learning rate warmup | 1700 | 1100 | 500 |
| Weight decay | 0.01 | 0.01 | 0.0001 |
| **Loss Function** | | | |
| Spatial loss factor | 40 | 20 | 1 |
| Spatial difference loss type | minus_log | minus_log | minus_log |
| **Pretraining-Specific Parameters** | | | |
| Number of hub ROIs masked | 250 | - | - |
| Temporal masking window size | 20 | - | - |
| Window interval rate | 4 | - | - |

**Supplementary Table S10. Final optimized hyperparameters for baseline models.** Hyperparameter values are reported for models trained on both low-pass filtered and unfiltered fMRI data. Where the optimal hyperparameter was the same for both conditions, a single value is shown. Where they differed, values are presented as Filtered / Unfiltered. Hyperparameters include optimizer type, learning rate, batch size, weight decay, learning rate scheduler, and model-specific parameters (e.g., kernel type for SVM, tree depth for XGBoost). **Abbreviations:** FC: functional connectivity, TS: time series, SVM: support vector machine.

| Hyperparameter | SVM | XGBoost | BNT | BolT | Brain LM | Vanilla BERT |
|---|---|---|---|---|---|---|
| **Library Version** | scikit-learn 1.4.0 | xgboost 2.0.3 | PyTorch 2.7.1 | PyTorch 2.7.1 | PyTorch 2.7.1 | Transformers 4.54.0 |
| **Input Data** | FC | FC | FC | TS | TS | TS |
| **Optimizer** | - | - | Adam | Adam | AdamW | AdamW |
| **Learning Rate** | - | 0.3 / 0.05 | 1.00E-04 | 1e-4 / 2e-4 | 1.00E-04 | 5.00E-05 |
| **Batch Size** | - | - | 32 / 8 | 16 | 64 | 16 |
| **Weight Decay** | - | - | 1.00E-03 | 1e-4 / 0 | 1.00E-05 | 1.00E-03 |
| **Scheduler** | - | - | Cosine | - | Linear | Step |
| **Kernel** | linear | - | - | - | - | - |
| **C** | 0.001 | - | - | - | - | - |
| **Max Depth** | - | 10 | - | - | - | - |
| **Min Child W.** | - | 1 / 4 | - | - | - | - |
| **Gamma** | - | 0.1 / 0.4 | - | - | - | - |
| **Colsample** | - | 0.9 / 0.8 | - | - | - | - |
| **LR Gamma** | - | - | - | - | - | 0.91 |
| **LR Step Size** | - | - | - | - | - | 4000 |
| **Pretrained Model** | - | - | - | - | 111M | - |

**Supplementary Table S11. Classification performance across subgroups.** Performance metrics for OCD vs. healthy control classification using the MBBN from-scratch model. Values represent mean ± standard deviation across 10 random seeds. The overall analysis (N = 1,706) excludes sites with TR > 2 seconds. Subgroups are based on medication status (medicated vs. unmedicated) and age groups (adults vs. pediatrics). **Abbreviations:** AUROC: area under the receiver operating characteristic curve, HC: healthy controls, OCD: obsessive-compulsive disorder, MBBN: Multi-Band Brain Network, Bal. Acc.: balanced accuracy, SD: standard deviation.

| Subgroup Analysis | N (OCD) | N (HC) | AUROC ± SD | Bal. Acc. ± SD | Sensitivity ± SD | Specificity ± SD |
|---|---|---|---|---|---|---|
| **Whole sample OCD vs. HC** | 869 | 837 | 65.26 ± 3.85 | 60.36 ± 4.12 | 66.86 ± 16.87 | 53.87 ± 12.47 |
| **Unmedicated OCD vs. HC** | 415 | 837 | 68.60 ± 3.69 | 62.01 ± 3.20 | 61.75 ± 18.32 | 62.27 ± 15.73 |
| **Medicated OCD vs. HC** | 447 | 837 | 69.65 ± 2.56 | 61.18 ± 4.95 | 43.04 ± 22.69 | 78.21 ± 15.15 |
| **Adults OCD vs. Adults HC** | 760 | 754 | 65.27 ± 3.55 | 58.22 ± 2.86 | 54.88 ± 22.04 | 61.56 ± 22.26 |
| **Pediatrics OCD vs. Pediatrics HC** | 109 | 83 | 54.42 ± 11.19 | 52.80 ± 8.82 | 70.93 ± 13.93 | 34.67 ± 14.90 |

**Supplementary Table S12. (From-scratch MBBN model) Full multiple linear regression results predicting log-transformed uncertainty.** The table shows standardized regression coefficients (β) for continuous predictors and unstandardized coefficients for binary/categorical predictors, along with 95% confidence intervals (CI) and p-values based on heteroscedasticity-consistent standard errors (HC3). Continuous predictors were z-scored before model fitting. Reference categories for categorical variables: Sex = "Male" (coded 0), Scanner = "GE". Binary QC variables coded as 0 = "Good", 1 = "Uncertain". "-" indicates that the predictor was not applicable/included in that specific model. Asterisks indicate significance levels: $p < .05$*, $p < .01$**, $p < .001$***. **Abbreviations:** HC: healthy controls, OCD: obsessive-compulsive disorder, ICA-AROMA: automatic removal of motion artifacts via independent component analysis, EPI: echo planar imaging, FD: framewise displacement, BOLD: blood-oxygen-level-dependent, Y-BOCS: Yale-Brown obsessive-compulsive scale, SNR: signal-to-noise ratio, T1w: T1-weighted.

| Predictor | Whole Sample β | Whole Sample 95% CI | Whole Sample *p* | HC-Only β | HC-Only 95% CI | HC-Only *p* | OCD-Only β | OCD-Only 95% CI | OCD-Only *p* |
|---|---|---|---|---|---|---|---|---|---|
| **Age** | -0.0100 | (-0.025, 0.004) | .174 | 0.0021 | (-0.019, 0.023) | .841 | -0.0359 | (-0.059, -0.013) | .002** |
| **Scanner: Philips** | -0.012 | (-0.057, 0.033) | .602 | -0.0336 | (-0.096, 0.029) | .290 | -0.1351 | (-0.206, -0.065) | < .001*** |
| **Scanner: Siemens** | 0.0686 | (0.033, 0.104) | < .001*** | 0.0259 | (-0.026, 0.078) | .332 | 0.0125 | (-0.041, 0.066) | .645 |
| **Mean FD (motion)** | 0.0304 | (0.017, 0.044) | < .001*** | 0.0215 | (0.000, 0.043) | .047* | 0.0383 | (0.018, 0.059) | < .001*** |
| **Sample size (site)** | -0.0802 | (-0.095, -0.065) | < .001*** | -0.0699 | (-0.094, -0.046) | < .001*** | -0.0658 | (-0.089, -0.042) | < .001*** |
| **Sex** | -0.0029 | (-0.031, 0.025) | .836 | 0.0088 | (-0.032, 0.050) | .676 | -0.0133 | (-0.053, 0.027) | .516 |
| **BOLD confounds QC** | 0.1111 | (0.071, 0.151) | < .001*** | 0.1321 | (0.071, 0.194) | < .001*** | 0.0478 | (-0.010, 0.105) | .104 |
| **EPI norm QC** | -0.0133 | (-0.067, 0.041) | .629 | -0.0142 | (-0.093, 0.065) | .723 | 0.0698 | (-0.011, 0.150) | .090 |
| **ICA-AROMA QC** | 0.0211 | (-0.016, 0.058) | .260 | -0.0146 | (-0.067, 0.038) | .583 | 0.0934 | (0.042, 0.145) | < .001*** |
| **Skull strip QC** | -0.0598 | (-0.156, 0.037) | .224 | -0.0272 | (-0.166, 0.112) | .701 | 0.0184 | (-0.097, 0.133) | .753 |
| **TSNR QC** | 0.0128 | (-0.021, 0.047) | .462 | 0.0063 | (-0.042, 0.055) | .798 | -0.0167 | (-0.066, 0.033) | .509 |
| **OCD** | -0.0079 | (-0.036, 0.020) | .575 | - | - | - | - | - | - |
| **Medication** | - | - | - | - | - | - | -0.0686 | (-0.113, -0.025) | .002** |
| **Y-BOCS score** | - | - | - | - | - | - | 0.0033 | (-0.019, 0.025) | .770 |
| **Age of onset** | - | - | - | - | - | - | -0.0057 | (-0.051, 0.039) | .802 |
| **Anxiety current** | - | - | - | - | - | - | -0.0382 | (-0.087, 0.011) | .129 |
| **Depression current** | - | - | - | - | - | - | -0.0153 | (-0.068, 0.038) | .570 |
| **No. Observations** | 1689 | | | 827 | | | 683 | | |
| **Df Residuals** | 1676 | | | 815 | | | 666 | | |
| **Df Model** | 12 | | | 11 | | | 16 | | |
| **R-squared** | .10 | | | .08 | | | .18 | | |
| **Adj. R-squared** | .09 | | | .06 | | | .16 | | |
| **F-statistic** | 17.05 | | | 6.02 | | | 10.4 | | |
| **Prob (F-statistic)** | < .001*** | | | < .001*** | | | < .001*** | | |
| **Log-Likelihood** | -276.47 | | | -143.29 | | | -47.191 | | |
| **AIC** | 578.9 | | | 310.6 | | | 128.4 | | |
| **BIC** | 649.6 | | | 367.2 | | | 205.3 | | |

**Supplementary Table S13. (Pretrained MBBN model) Full multiple linear regression results predicting log-transformed uncertainty.** The table shows standardized regression coefficients (β) for continuous predictors and unstandardized coefficients for binary/categorical predictors, along with 95% confidence intervals (CI) and p-values based on heteroscedasticity-consistent standard errors (HC3). Continuous predictors were z-scored before model fitting. Reference categories for categorical variables: Sex = "Male" (coded 0), Scanner = "GE". Binary QC variables coded as 0 = "Good", 1 = "Uncertain". "-" indicates that the predictor was not applicable/included in that specific model. Asterisks indicate significance levels: $p < .05$*, $p < .01$**, $p < .001$***. **Abbreviations:** HC: healthy controls, OCD: obsessive-compulsive disorder patients, ICA-AROMA: automatic removal of motion artifacts via independent component analysis, EPI: echo planar imaging, FD: framewise displacement, BOLD: blood-oxygen-level-dependent, Y-BOCS: Yale-Brown obsessive-compulsive scale, SNR: signal-to-noise ratio, T1w: T1-weighted.

| Predictor | Whole Sample β | Whole Sample 95% CI | Whole Sample *p* | HC-Only β | HC-Only 95% CI | HC-Only *p* | OCD-Only β | OCD-Only 95% CI | OCD-Only *p* |
|---|---|---|---|---|---|---|---|---|---|
| **Age** | -0.0152 | (-0.029, -0.001) | .032* | 0.0020 | (-0.017, 0.021) | .831 | -0.0410 | (-0.063, -0.019) | < .001*** |
| **Scanner: Philips** | 0.0300 | (-0.014, 0.074) | .185 | 0.0209 | (-0.042, 0.084) | .518 | -0.1250 | (-0.187, -0.063) | < .001*** |
| **Scanner: Siemens** | 0.1770 | (0.144, 0.210) | < .001*** | 0.1705 | (0.125, 0.216) | < .001*** | 0.0953 | (0.044, 0.146) | < .001*** |
| **Mean FD (motion)** | 0.0328 | (0.019, 0.047) | < .001*** | 0.0208 | (0.001, 0.041) | .039* | 0.0563 | (0.035, 0.078) | < .001*** |
| **Sample size (site)** | -0.1195 | (-0.134, -0.105) | < .001*** | -0.1106 | (-0.131, -0.090) | < .001*** | -0.1154 | (-0.140, -0.090) | < .001*** |
| **Sex** | 0.0090 | (-0.018, 0.036) | .509 | 0.0273 | (-0.011, 0.066) | .163 | -0.0116 | (-0.050, 0.027) | .559 |
| **BOLD confounds QC** | 0.1134 | (0.075, 0.152) | < .001*** | 0.1305 | (0.076, 0.185) | < .001*** | 0.0291 | (-0.030, 0.088) | .337 |
| **EPI norm QC** | 0.0021 | (-0.052, 0.056) | .939 | 0.0190 | (-0.060, 0.098) | .635 | 0.0838 | (-0.004, 0.172) | .062 |
| **ICA-AROMA QC** | 0.0402 | (0.006, 0.075) | .022* | 0.0338 | (-0.015, 0.083) | .175 | 0.0854 | (0.037, 0.134) | < .001*** |
| **Skull strip QC** | -0.1291 | (-0.238, -0.021) | .020* | -0.0762 | (-0.232, 0.079) | .337 | -0.0705 | (-0.221, 0.080) | .357 |
| **TSNR QC** | 0.0397 | (0.005, 0.074) | .024* | 0.0304 | (-0.018, 0.079) | .217 | 0.0019 | (-0.049, 0.052) | .943 |
| **OCD** | 0.0091 | (-0.017, 0.035) | .500 | - | - | - | - | - | - |
| **Medication** | - | - | - | - | - | - | -0.0916 | (-0.136, -0.048) | < .001*** |
| **Y-BOCS score** | - | - | - | - | - | - | 0.0139 | (-0.006, 0.034) | .177 |
| **Age of onset** | - | - | - | - | - | - | -0.0098 | (-0.053, 0.033) | .655 |
| **Anxiety current** | - | - | - | - | - | - | -0.0256 | (-0.076, 0.025) | .317 |
| **Depression current** | - | - | - | - | - | - | 0.0107 | (-0.042, 0.064) | .693 |
| **No. Observations** | 1689 | | | 827 | | | 683 | | |
| **Df Residuals** | 1676 | | | 815 | | | 666 | | |
| **Df Model** | 12 | | | 11 | | | 16 | | |
| **R-squared** | .20 | | | .17 | | | .28 | | |
| **Adj. R-squared** | .19 | | | .16 | | | .26 | | |
| **F-statistic** | 38.91 | | | 17.2 | | | 18.72 | | |
| **Prob (F-statistic)** | < .001*** | | | < .001*** | | | < .001*** | | |
| **Log-Likelihood** | -209.64 | | | -83.943 | | | -27.748 | | |
| **AIC** | 445.3 | | | 191.9 | | | 89.5 | | |
| **BIC** | 515.9 | | | 248.5 | | | 166.4 | | |
